# Evaluating multiple next-generation sequencing derived tumor features to accurately predict DNA mismatch repair status

**DOI:** 10.1101/2022.06.20.22276419

**Authors:** Romy Walker, Peter Georgeson, Khalid Mahmood, Jihoon E. Joo, Enes Makalic, Mark Clendenning, Julia Como, Susan Preston, Sharelle Joseland, Bernard J. Pope, Ryan Hutchinson, Kais Kasem, Michael D. Walsh, Finlay A. Macrae, Aung K. Win, John L. Hopper, Dmitri Mouradov, Peter Gibbs, Oliver M. Sieber, Dylan E. O’Sullivan, Darren R. Brenner, Steven Gallinger, Mark A. Jenkins, Christophe Rosty, Ingrid M. Winship, Daniel D. Buchanan

## Abstract

Identifying tumor DNA mismatch repair deficiency (dMMR) is important for precision medicine. We assessed tumor features, individually and in combination, in whole-exome sequenced (WES) colorectal cancers (CRCs) and in panel sequenced CRCs, endometrial cancers (ECs) and sebaceous skin tumors (SSTs) for their accuracy in detecting dMMR. CRCs (n=300) with WES, where MMR status was determined by immunohistochemistry, were assessed for microsatellite instability (MSMuTect, MANTIS, MSIseq, MSISensor), COSMIC tumor mutational signatures (TMS) and somatic mutation counts. A 10-fold cross-validation approach (100 repeats) evaluated the dMMR prediction accuracy for 1) individual features, 2) Lasso statistical model and 3) an additive feature combination approach. Panel sequenced tumors (29 CRCs, 22 ECs, 20 SSTs) were assessed for the top performing dMMR predicting features/models using these three approaches. For WES CRCs, 10 features provided >80% dMMR prediction accuracy, with MSMuTect, MSIseq, and MANTIS achieving ≥99% accuracy. The Lasso model achieved 98.3%. The additive feature approach with ≥3/6 of MSMuTect, MANTIS, MSIseq, MSISensor, INDEL count or TMS ID2+ID7 achieved 99.7% accuracy. For the panel sequenced tumors, the additive feature combination approach of ≥3/6 achieved accuracies of 100%, 95.5% and 100%, for CRCs, ECs, and SSTs, respectively. The microsatellite instability calling tools performed well in WES CRCs, however, an approach combining tumor features may improve dMMR prediction in both WES and panel sequenced data across tissue types.

## Introduction

DNA mismatch-repair (MMR) deficiency (dMMR) is an important molecular phenotype of solid tumors characterized by the presence of microsatellite instability (MSI) and/or loss of expression of one or more of the DNA MMR proteins, MLH1, MSH2, MSH6 and PMS2. Identifying dMMR tumors is important for understanding disease prognosis^1^, response to immune checkpoint inhibition therapy^2^ and to identify people with Lynch syndrome. Lynch syndrome is the most common inherited cancer predisposition disorder and, therefore, the Evaluation of Genomic Applications in Practice and Prevention Working Group recommends that all newly diagnosed colorectal (CRC) and endometrial cancers (EC) are screened for dMMR to improve the identification of carriers^3, 4^.

The dMMR mutator phenotype arises in tumors where errors occur during the DNA replication process^5^. Specifically, defects in the components of the MMR system responsible for the recognition of mismatches such as single nucleotide variants (SNVs) and insertion-deletions (INDELs), can lead to the development of numerous frameshift mutations in coding and non-coding microsatellite regions^6^. dMMR is related to biallelic inactivation of one of the MMR genes, resulting from either somatic methylation of the *MLH1* gene promoter region^7^ or double somatic MMR gene mutations^8^ (sporadic dMMR), or germline pathogenic variants in the MMR genes^9^ or deletions in the 3′ end of the *EPCAM* gene^10^ (inherited dMMR). CRC, EC and sebaceous skin tumors (SSTs), including sebaceous adenomas, carcinomas and sebaceomas, are tissue types that demonstrate the highest frequencies of dMMR where up to 26%^11^, 31%^11^ and 31%^12^ of these tissue types, respectively, present with the dMMR phenotype, followed by stomach cancer at 19%^11^.

The most common approach for identifying dMMR tumors is by assessing MMR protein expression through immunohistochemistry (MMR IHC)^13, 14^ and/or by testing for high levels of microsatellite instability using polymerase chain reactions (MSI-PCR)^15^. While both screening methodologies are commonly used, each present advantages and limitations. The advantages of performing MMR IHC include simple experimental execution, short turnaround time, low associated costs as well as giving an indication of the defective gene^16^. However, false positive or false negative MMR IHC results can occur due to technical artefacts, variable performance of different MMR antibodies and inherent variability in the interpretation of the staining by different pathologists^16^. Further challenges include the interpretation of weaker staining in less proliferative tissue and heterogenous patterns of MMR protein loss^17–24^.

While MMR IHC is more widely adopted in the clinical setting, MSI-PCR remains the gold standard for detecting dMMR^16^; to date multiple markers have been identified to call MSI in tumor samples^25^. The limitations for MSI-PCRs include additional laboratory implementation requirements related to tissue DNA extraction and increased labor costs; both can lead to a delay in receiving test results^16^. Nonetheless, MMR IHC and MSI-PCR methodologies have proven to be effective for identifying dMMR in CRC samples^26^ with a reported concordance of 91.9%^16^, but the accuracy for either of these tools can decrease when applied to different tissue types^27^. As next-generation sequencing (NGS) becomes more widely adopted for precision oncology, there is an increasing need to accurately determine tumor MMR status using NGS data.

To date, several tools have been developed to assess MSI from NGS data, including MSISensor^28^, MSIseq^29^, MANTIS^30^ and more recently MSMuTect^31^. To the best of our knowledge, the comparison of these four MSI tools on the same tumors has not yet been performed. In addition to MSI, other tumor features derived from NGS have been shown to be associated with dMMR, such as tumor mutational burden (TMB)^32^ and tumor mutational signatures (TMS)^33^. TMB, characterized by high SNV and INDEL counts, is a biomarker for response to immune checkpoint inhibition therapy^34, 35^ and is increased in dMMR tumors^36^.

TMS aggregate tens to thousands of the observed somatic mutations within a tumor into patterns related to the underlying mutational processes^37, 38^. The predominant TMS framework, published on the COSMIC website, defines 107 different signature definitions categorized into three distinct subgroups: 1) 78 single base substitutions (SBS) where seven of the SBS signatures (SBS6, SBS14, SBS15, SBS20, SBS21, SBS26 and SBS44) are associated with dMMR; 2) 18 small (1 to 50 base pair) insertions and deletions or ID signatures where ID1, ID2, and ID7 are associated with dMMR, and 3) 11 doublet base substitutions or DBS signatures where DBS7 and DBS10 have both been previously associated with dMMR^33^. However, DBS signatures have a reported low prevalence in CRC compared with other tissue types so were excluded from our study^38^. Previously, we have shown that the combination of individual TMS can improve the ability of TMS to discriminate important molecular and genetic subtypes of CRC, including identifying germline biallelic carriers of pathogenic variants in the *MUTYH* gene by combining SBS18 and SBS36^39, 40^. We further observed that the combination of ID2 with ID7 (TMS ID2+ID7) was the most informative for differentiating dMMR from pMMR CRCs amongst all possible TMS combinations^39^. To date, the comparison of MSI calling tools, somatic mutation counts, TMB and TMS tumor features for determining the dMMR status in CRC tumors has not yet been undertaken.

In this study, we assessed 104 tumor features derived from whole-exome sequencing (WES) (**Table 1**), consisting of the MSI prediction tools (MSMuTect, MANTIS, MSIseq and MSISensor), TMS (78 SBS and 18 ID signatures), TMS ID2+ID7, TMB and individual SNV and INDEL somatic mutation counts for their accuracy in predicting dMMR status in 300 well-characterized CRCs. Secondly, we investigated whether a combination of these tumor features, using either a statistical model or a simple approach that added individual features together (additive feature combination), could improve the dMMR prediction accuracy in WES CRC tumors. Finally, we evaluated the effectiveness of the top performing tumor features from the WES analysis, individually and in combination, in an independent set of CRC, EC and SST tumors that had undergone targeted multigene panel sequencing for their dMMR prediction accuracy.

**Table 1.**
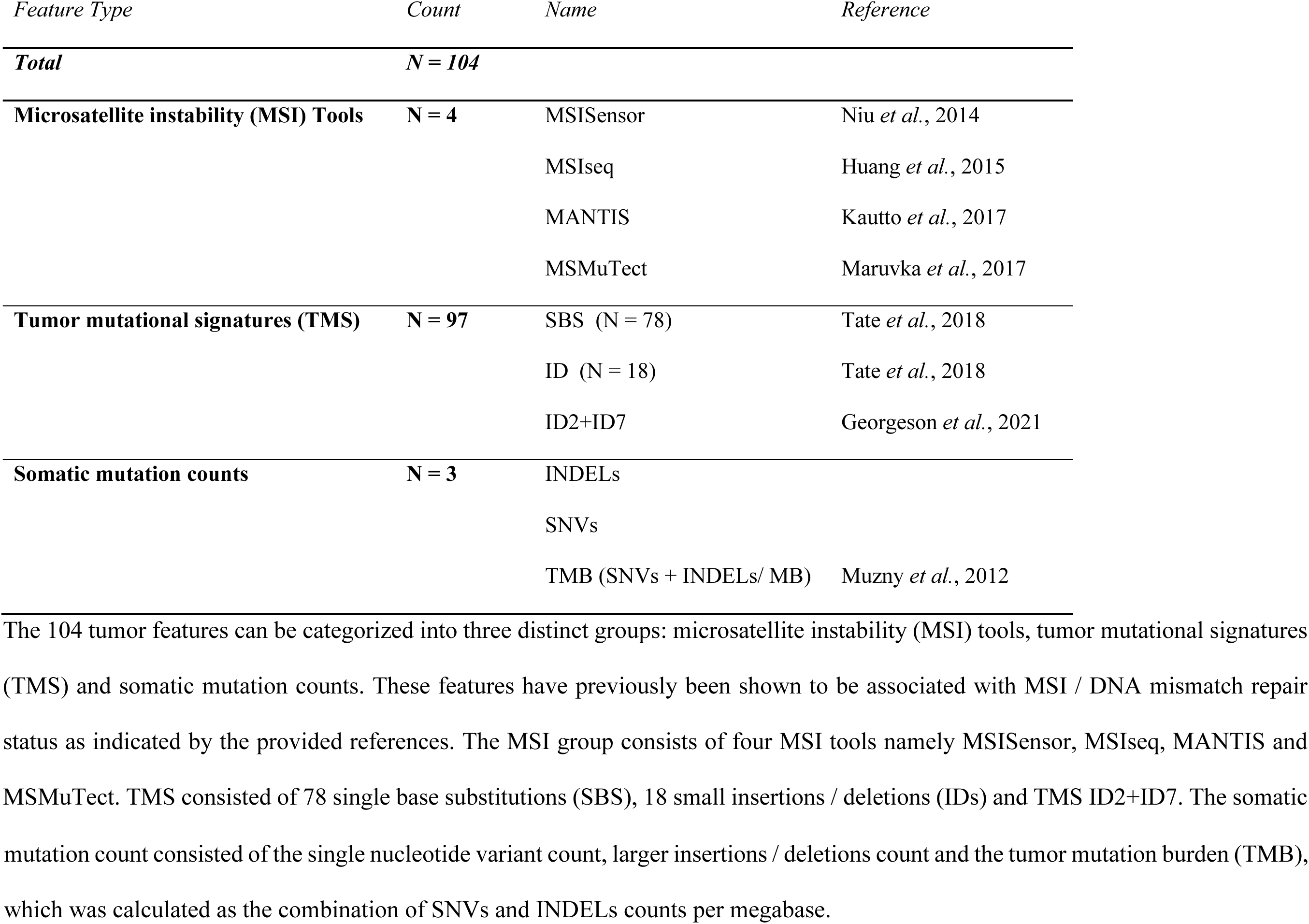
The breakdown of the 104 tumor features calculated from next generation sequencing analysis included in this study.

| *Feature Type* | *Count* | *Name* | *Reference* |
| --- | --- | --- | --- |
| ***Total*** | ***N = 104*** |  |  |
| **Microsatellite instability (MSI) Tools** | **N = 4** | MSISensor | Niu *et al.*, 2014 |
|  |  | MSIseq | Huang *et al.*, 2015 |
|  |  | MANTIS | Kautto *et al.*, 2017 |
|  |  | MSMuTect | Maruvka *et al.*, 2017 |
| **Tumor mutational signatures (TMS)** | **N = 97** | SBS (N = 78) | Tate *et al.*, 2018 |
|  |  | ID (N = 18) | Tate *et al.*, 2018 |
|  |  | ID2+ID7 | Georgeson *et al.*, 2021 |
| **Somatic mutation counts** | **N = 3** | INDELs |  |
|  |  | SNVs |  |
|  |  | TMB (SNVs + INDELs/ MB) | Muzny *et al.*, 2012 |

The 104 tumor features can be categorized into three distinct groups: microsatellite instability (MSI) tools, tumor mutational signatures (TMS) and somatic mutation counts. These features have previously been shown to be associated with MSI / DNA mismatch repair status as indicated by the provided references. The MSI group consists of four MSI tools namely MSISensor, MSIseq, MANTIS and MSMuTect. TMS consisted of 78 single base substitutions (SBS), 18 small insertions / deletions (IDs) and TMS ID2+ID7. The somatic
mutation count consisted of the single nucleotide variant count, larger insertions / deletions count and the tumor mutation burden (TMB), which was calculated as the combination of SNVs and INDELs counts per megabase.

## Materials and Methods

### Study Cohort

The study population included men and women retrospectively identified from five studies where pMMR or dMMR status was determined by MMR IHC and where an etiology for dMMR status could be defined, namely a sporadic etiology caused by tumor *MLH1* methylation or double somatic MMR mutations, or an inherited etiology caused by a germline MMR gene pathogenic variant (Lynch syndrome). The breakdown of participants included in this study by their dMMR and pMMR status, tissue type and by WES or panel sequencing is shown in **Figure 1**:

**1)** the ANGELS study (*<u>A</u>pplying <u>N</u>ovel <u>G</u>enomic approaches to <u>E</u>arly-onset and suspected <u>L</u>ynch <u>S</u>yndrome colorectal and endometrial cancers*)^39^ recruited participants that were diagnosed with CRC or EC between 2014 – 2021 who were referred from family cancer clinics across Australia (n=79). All ANGELS study participants provided informed consent and the study was approved by the University of Melbourne human research ethics committee (HREC#1750748) and institutional review boards at each family cancer clinic;
**2)** CRC- or EC-affected participants from the ACCFR (*<u>A</u>ustralasian <u>C</u>olorectal <u>C</u>ancer <u>F</u>amily <u>R</u>egistry*) were selected from both population-based and clinic-based recruitment (n=139);
**3)** CRC-affected participants from the OFCCR (*<u>O</u>ntario <u>F</u>amilial <u>C</u>olorectal <u>C</u>ancer <u>R</u>egistry*) were population-based patients (<50 years old) recruited from the Cancer Care Ontario, Toronto, Canada (n=53). Study participants from both the ACCFR and OFCCR were recruited between 1998 and 2008, and were included according to the recruitment policy and eligibility criteria previously described^41, 42^. Informed consent was obtained from all study participants and the study protocol was approved by the institutional human ethics committee at both study sites;
**4)** CRC-affected participants from the WEHI study (*<u>W</u>alter and <u>E</u>liza <u>H</u>all <u>I</u>nstitute of Medical Research*) were recruited from the Royal Melbourne Hospital (Parkville, VIC, Australia) and the Western Hospital Footscray (Footscray, VIC, Australia), between Jan 1, 1993, and Dec 31, 2009^39^. All patients provided written informed consent. The study was approved by human research ethics committees at both sites (HREC 12/19) (n = 80);
**5)** SST-affected participants from the MTS study (*<u>M</u>uir-<u>T</u>orre <u>S</u>yndrome Study*) were referred between July 2016 and September 2021 following clinical diagnostic MMR IHC testing by Sullivan Nicolaides Pathology service in Brisbane^12^ or by family cancer clinics in Australia. Informed consent was obtained from the study participants and the study protocol was approved by the human research ethics committee from the University of Melbourne (HREC#1648355) and by the relevant institutional human ethics committees (n = 20).

**Figure 1.**
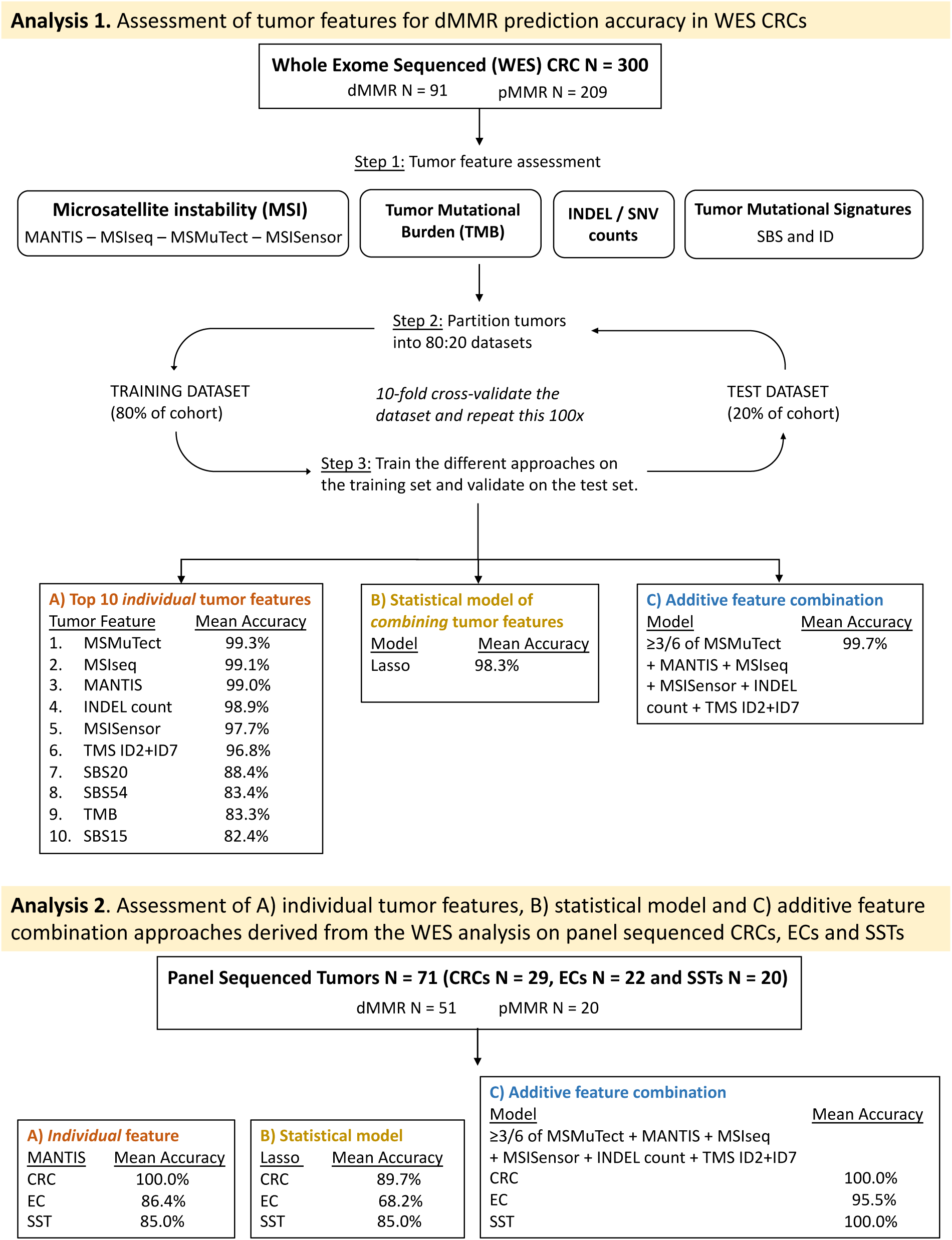
Overview of the study design. In total, 300 whole-exome sequenced (WES) colorectal cancers (CRCs) consisting of 91 DNA mismatch repair deficient (dMMR) and 209 DNA mismatch repair proficient (pMMR) tumors were analyzed. We investigated 104 tumor features for their ability to distinguish dMMR from pMMR tumors consisting of four MSI tools, 97 tumor mutational signature definitions (TMS), tumor mutation burden (TMB) calculated as mutations per mega base, somatic insertion / deletion (INDEL) and somatic single nucleotide variant (SNV) counts. We performed a 10-fold cross-validation approach with 100 repeats to calculate the mean accuracy on the test dataset. (A) The top 10 ranked individual tumor features, (B) a Lasso regression model and (C) an additive feature combination approach was tested to determine the benefit of combining tumor features to improve dMMR prediction. The findings from these three approaches were tested on an independent set of targeted panel sequenced tumors of CRC, endometrial cancer (EC) and sebaceous skin tumor (SST) tissue types with reported mean accuracies.

### Tumor Categorization

MMR IHC testing was performed on formalin-fixed paraffin embedded (FFPE) tissues for all four MMR proteins for the ACCFR and OFCCR as previously described^42–44^, and a subset of these tumors also underwent MSI-PCR testing as previously described^45^. MMR IHC testing for the ANGELS and MTS studies was part of routine clinical assessment in pathology laboratories across Australia, reported by the duty pathologist. Fresh-frozen tissue specimens from the WEHI study were assessed for MLH1, MSH2 and MSH6 MMR IHC and MSI-PCR tested using BAT25, BAT26, D5S346, D2S123 and D17S250 MSI markers. Germline MMR gene testing (as described in Buchanan *et al.*^43^) and tumor *MLH1* promoter methylation testing by MethyLight (as described in Buchanan *et al.*^46^) were performed on all dMMR tumors showing loss of MLH1/PMS2 protein expression or sole PMS2 loss by IHC. Tumors were considered to have double somatic MMR mutations when they were found to have two pathogenic/likely pathogenic somatic mutations or a single somatic pathogenic/likely pathogenic mutation in combination with presence of loss of heterozygosity. Germline pathogenic variants and somatic MMR gene mutations were confirmed in WES and targeted panel sequencing data prior to analysis. Therefore, for each of the dMMR tumors included in this study we could confirm an inherited or acquired cause for their respective pattern of MMR IHC protein loss. Concurrently, for the pMMR tumors, we did not find evidence of a germline MMR pathogenic variant or double MMR somatic mutation in these tumor samples.

All tumors in the study were assigned to one of four categories based on dMMR or pMMR status determined from MMR IHC and/or MSI-PCR and based on the cause for dMMR:

**1) dMMR-Lynch syndrome (dMMR-LS)** – identified carrier of a germline pathogenic variant in one of the DNA MMR genes where the corresponding tumor showed commensurate loss of MMR protein expression by IHC;
**2) dMMR-*MLH1* methylation (dMMR-MLH1me)** – tumors were positive for methylation of the *MLH1* gene promoter “C region”^47^ and showed loss of MLH1 and PMS2 protein expression by IHC without a germline MMR gene pathogenic variant;
**3) dMMR-double somatic (dMMR-DS)** – tumors harbored two somatic mutations (SNVs and/or loss of heterozygosity) in the same MMR gene that showed loss of protein expression by IHC with no identified pathogenic germline MMR gene variant; and
**4) MMR-proficient** (**pMMR**) – tumors showed normal expression of all four MMR proteins and did not show presence of double somatic MMR gene mutations or a germline MMR gene pathogenic variant.

The three dMMR subtypes dMMR-LS, dMMR-DS and dMMR-MLH1me were combined as a single dMMR tumor group in downstream analysis.

### Whole-Exome and Targeted Panel Sequencing Capture Regions

The targeted panel was based on the design described in Zaidi *et al*.^48^ consisting of probes targeting the following regions: 1) 298 genes incorporating key hereditary CRC^49–51^ and EC^52^ risk genes and genes that are frequently mutated as identified by The Cancer Genome Atlas (TCGA) data^32, 53, 54^, 2) 28 microsatellite loci including the five ‘gold standard’ MSI markers (BAT25, BAT26, NR-21, NR-24, and MONO-27) currently implemented in routine MSI-PCR diagnostics, 3) 212 homopolymer regions distributed genome-wide to assess for MSI in tumor samples and 4) 56 copy number variants known to be susceptible to copy number changes in CRCs. The panel capture was 2.005 megabases (Mb) in size. The WES capture incorporates all exonic regions within the genome and is 67.296 Mb in size. The panel additionally included capture of intronic regions within the MMR genes, which the WES capture did not cover.

### Next-Generation Sequencing

In total, 300 CRC tumors were sequenced by WES and 71 tumors (29 CRCs, 22 ECs and 20 SSTs) were sequenced by the targeted multigene panel (**Figure 1**). FFPE CRC, EC or SST tissues were macrodissected and DNA extracted using the QIAmp DNA FFPE Tissue Kit (Qiagen, Hilden, Germany) according to the manufacturer’s instructions. Peripheral blood-derived DNA was extracted using the DNeasy blood and tissue kit (Qiagen, Hilden, Germany) and sequenced as germline references.

The WES capture was the Agilent Clinical Research Exome V2 kit (Agilent Technologies Santa Clara, United States) with sequencing performed on an Illumina NovaSeq 6000 comprising 150 base pair (bp) paired-end reads performed at the Australian Genome Research Facility^39^. For the WEHI CRCs, exome-enrichment was performed using the TruSeq Exome Enrichment Kit (Illumina, San Diego, United States) and 100 bp paired-end read sequencing performed on an Illumina HiSeq 2000 at the Australian Genome Research Facility^39^. The on-target coverage for the 300 WES samples had a median of 323.7 for the FFPE tumor DNA samples and 137.4 for blood-derived DNA samples, with an interquartile range of 111.8 – 426.4 and 100.6 – 204.9, respectively. Library preparation for targeted panel sequencing was performed using the SureSelect^TM^

Low Input Target Enrichment System (Agilent Technologies, Santa Clara, United States) using standard protocol and sequenced on an Illumina NovaSeq 6000 comprising 150 bp paired end reads performed at the Australian Genome Research Facility. The on-target coverage for the 71 panel sequenced samples was (median and interquartile range) 919.3 and 694.6 – 1164.9 for FFPE tumor DNA samples and 160.6 and 135.8 – 178.0 for blood-derived DNA samples.

### Bioinformatics Pipeline

For both WES and targeted panel sequenced samples, adapter sequences were trimmed from raw FASTQ files using trimmomatic 0.38^55^ and aligned to the GRCh37 human reference genome using Burrows-Wheeler Aligner v. 0.7.12. Germline variants, somatic variants (SNVs) and somatic INDELs were called using Strelka (v. 2.9.2., Illumina) using the recommended workflow^56^. TMS were calculated using the pre-defined set of 78 SBS and 18 ID signatures published on COSMIC as version 3.2 (COSMIC, https://cancer.sanger.ac.uk/signatures/, last accessed date: June 15, 2022)^33^. Variants outside the WES and panel capture regions were excluded and variants with the PASS filter called from Strelka were retained. Additional variant filters included were restrictions to a minimum depth of 50x for germline and tumor samples with a minimum variant allele frequency of 10% as detailed previously^39^.

### Selection of Features of Interest

The 104 tumor features selected for analysis in this study are shown in **Table 1**. Several tools have been developed to assess MSI from NGS data. Our analysis focused on MSMuTect^31^, MANTIS^30^, MSIseq^29^ and MSISensor^28^. Tumors were classified as having high levels of MSI (MSI-H) or as microsatellite stable (MSS). We assessed all SBS (n=78) and ID (n=18) TMS as described by COSMIC^33^, but the DBS TMS were excluded due to their reported low prevalence in CRCs^38^. Previously, we have shown that combining ID2 and ID7 TMS enabled detection of dMMR CRCs^39^ and, therefore, was included as a tumor feature in this study. Somatic mutation counts, namely SNVs or INDELs, as well as TMB (SNV and INDEL mutation count combined / Mb) were each included, given previous associations with tumor dMMR status^57^.

### Feature Performance Evaluation in WES data from CRCs

We assessed the 104 tumor features calculated from WES from 209 pMMR CRCs and 91 dMMR CRCs (pMMR:dMMR ratio = 2.3:1) (**Figure 1**). The dMMR CRCs comprised dMMR-LS tumors (n=49), dMMR-*MLH1*me tumors (n=26) and dMMR-DS tumors (n=16). All 300 CRCs were randomly partitioned into a training set (80% of CRCs) and a test set (20% of CRCs), while maintaining the same pMMR:dMMR ratio, using *caret* R package^58^. We performed a 10-fold cross validation approach on the training set (repeated 100x) to calculate the average classification accuracy by fitting a generalized linear model and determining the error rate, specificity, sensitivity, and the area under the curve (AUC) with corresponding 95% confidence intervals (CIs). Based on the unequal distribution of dMMR and pMMR tumors in the WES dataset, the *no information rate* was 69.5%, indicating that any feature with this prediction accuracy was equivalent to selecting a dMMR sample by chance.

Tumor feature analysis of the WES CRC dataset comprised of three different approaches:

#### A) Individual tumor feature assessment

Each of the 104 tumor features were assessed individually and then ranked by their accuracy in identifying dMMR tumors. Individual CRC tumor features with a prediction accuracy >80% from the WES data were considered good predictors for differentiating dMMR from pMMR tumors and were included in downstream analyses.

#### B) Generation of a statistical model by combining tumor features

We investigated whether combining tumor features using a Lasso penalized regression model^59^ could improve the overall dMMR prediction accuracy in CRC. Lasso enables the simultaneous parameter estimation and variable selection as well as having been shown to reduce overfitting when compared to conventional maximum likelihood regression models. Lasso regression has a tuning parameter called lambda that controls which features are included in the regression model by shrinking the coefficient or “weighting” of individual features within the model towards zero, helping with the exclusion of some of the features from integration into the final model via a penalization process using cross-validation.

#### C) Applying an additive feature combination count

Our third approach investigated combining the top ranked individual tumor features in an additive approach (additive feature combination). Specifically, the tumor features that achieved a mean prediction accuracy >95% from the WES CRC analysis (from part A), were included in this approach and added together to give an overall count. The bimodal distribution supported a majority vote decision on dMMR status.

### Assessment of individual tumor features, the statistical model and additive feature combination approaches derived from the WES analysis on panel sequenced CRCs, ECs, and SSTs

The top individual tumor features determined from (A), best performing Lasso model (B) and the additive feature combination approach (C) were then assessed for their dMMR prediction accuracy in three independent tumor sets comprised of n=29 CRCs, n=22 ECs and n=20 SSTs tested by targeted multigene panel sequencing. The *no information rate* for features analyzed from the panel dataset was at 71.8%, indicating a prediction accuracy of this value was similar to selecting a dMMR sample by chance.

### Statistical Analysis

All statistical analyses were done using the R programming language (v.4.1.0). The *tidyverse* package (v.1.3.1.)^60^ was used for data import, tidying and visualization purposes and the *caret* (v.6.0-9.0) package^58^ was used for cross-validation. Receiving operator curves (ROC) were generated using the *pROC* package (v.1.18.0)^61^, with the AUC being determined using the *cvAUC* package (v.1.1.4)^62^. Statistical models were fitted using the Lasso (*glmnet*, v.4.1-3)^63^ package. We used the *cutpointr* (v.1.1.1) package^64^ for estimation of the best “cut points” or “thresholds” which maximize the Youden-index (true positive rate minus false positive rate over all possible cut points), defined as the most optimal threshold in binary disease classification tasks. Here, the *cutpointr* package determines a recommended threshold that best differentiates dMMR from pMMR cases for each feature and validates its performance using bootstrapping. The average weight for each group was calculated using the *plyr* (v.1.0.7) package^65^. The *ggplot2* (v.3.3.5) package^66^ was used for data visualization in combination with *hrbrthemes* (v.0.8.0)^67^ for histogram generation and *ggrepel* (v.0.9.1)^68^ for histogram annotations. Correlation scores between the dMMR and pMMR groups were estimated by a *heteroscedastic two-tailed t-test*. P-values <0.05 were considered statistically significant. The 95% CIs for the WES data were calculated using the binomial (Clopper-Pearson) “exact” method^69^ and for the targeted panel data using the *binom* (v.1.1-1) package^70^ in R.

## Results

For the initial performance evaluation of 104 tumor features we assessed 209 (69.7%) pMMR CRCs and 91 (30.3%) dMMR CRCs sequenced by WES. The clinicopathological characteristics, pattern of MMR IHC loss and dMMR etiology are summarized in **Supplementary Table 1.** The mean age at CRC diagnosis (± standard deviation, SD) for the dMMR group was 51 ± 15.0 with 62.6% being female and 49 ± 16.3 with 55.5% being female for the pMMR group. The clinicopathological characteristics, pattern of MMR IHC loss and dMMR etiology for panel sequenced CRC (n=29), EC (n=22) and SST (n=20) tumors are summarized in **Supplementary Table 2**. Within the panel sequenced tumors, the proportion of dMMR for the CRC, EC and SST subsets was 72.4% (21/29), 81.8% (18/22) and 65.0% (13/20), respectively. The predominant dMMR subtype across the CRC WES and targeted panel sequenced tumors was dMMR-LS (53.8% and 66.7%, respectively). Within the dMMR subgroup, the most predominant pattern of loss observed in CRCs and ECs was MLH1/PMS2 (WES CRCs: 65.9%, panel CRCs: 47.6% and ECs: 50.0%), whereas for the SSTs tumors, this was MSH2/MSH6 loss (76.9%). Tumors showing less common patterns of MMR loss including solitary loss of MSH6 or PMS2 by IHC were present in both the WES CRCs (16.5%) and panel sequenced tumors (19.2%), however, sole PMS2 loss cases were absent from the EC and SST cohorts.

### Assessment of Tumor Features for dMMR Prediction Accuracy in WES CRCs

#### A) Individual tumor feature assessment

Twelve of the 104 tumor features derived from WES had a mean dMMR prediction accuracy >80% on the test dataset (**Table 2**). The mean accuracy for the remaining 92 features is shown in **Supplementary Table 3.** The four MSI tools were among the best predictors, with MSMuTect, MSIseq and MANTIS each achieving a mean prediction accuracy of ≥99.0% with MSMuTect achieving the highest accuracy (99.3%, 95% CI: 99.1%-99.5%) (**Table 2**). The combination of TMS ID2+ID7 achieved an accuracy of 96.8% (95% CI: 96.4%-97.2%), and outperformed these signatures individually (**Table 2**). To avoid collinearity issues between the combined TMS ID2+ID7 variable with the individual TMS ID2 and TMS ID7 features, the latter were excluded from downstream analysis as they provided a lower prediction score. Therefore, the remaining 10 features were considered as the top 10 dMMR predictors and included in subsequent analyses (**Figure 1**).

**Table 2.**
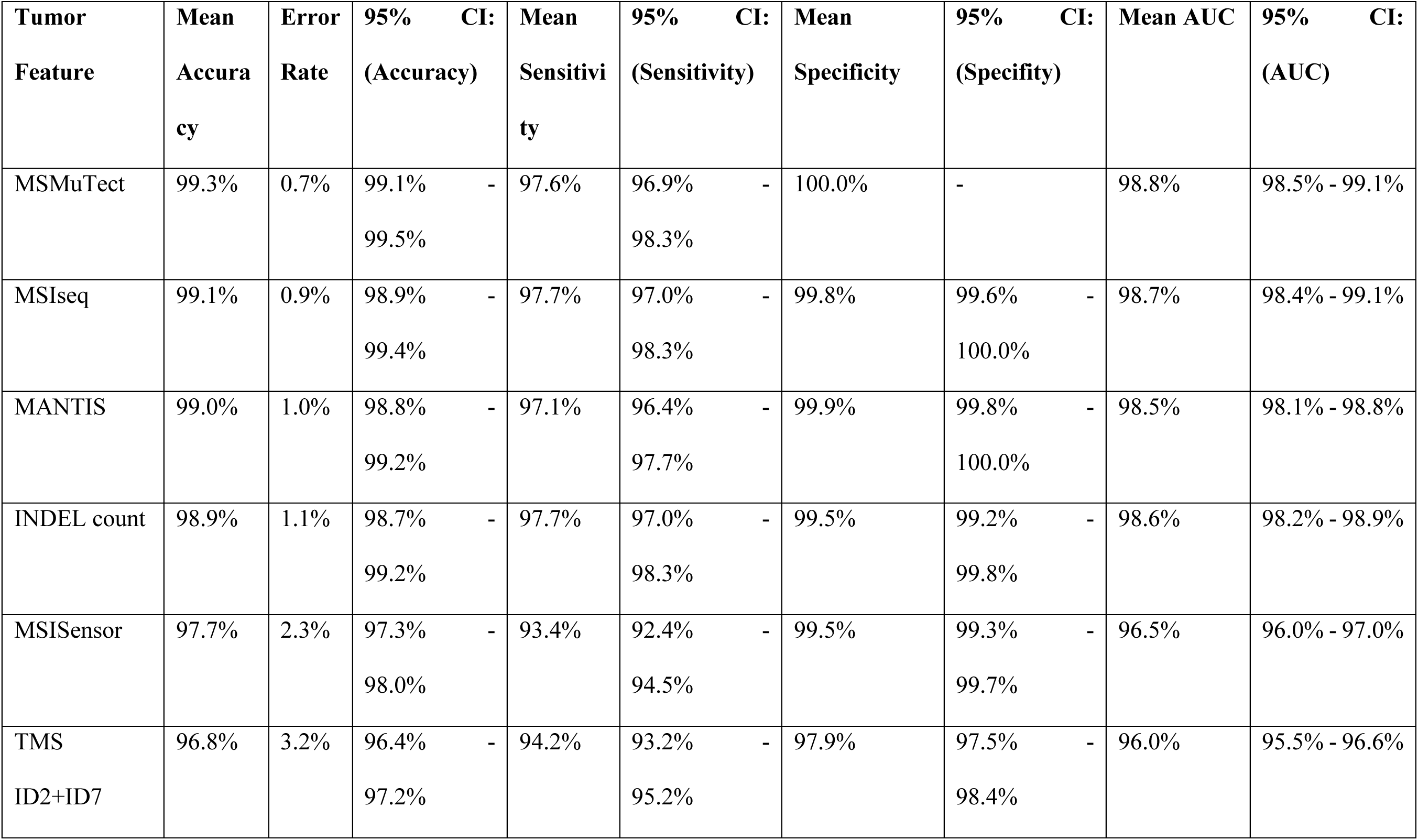
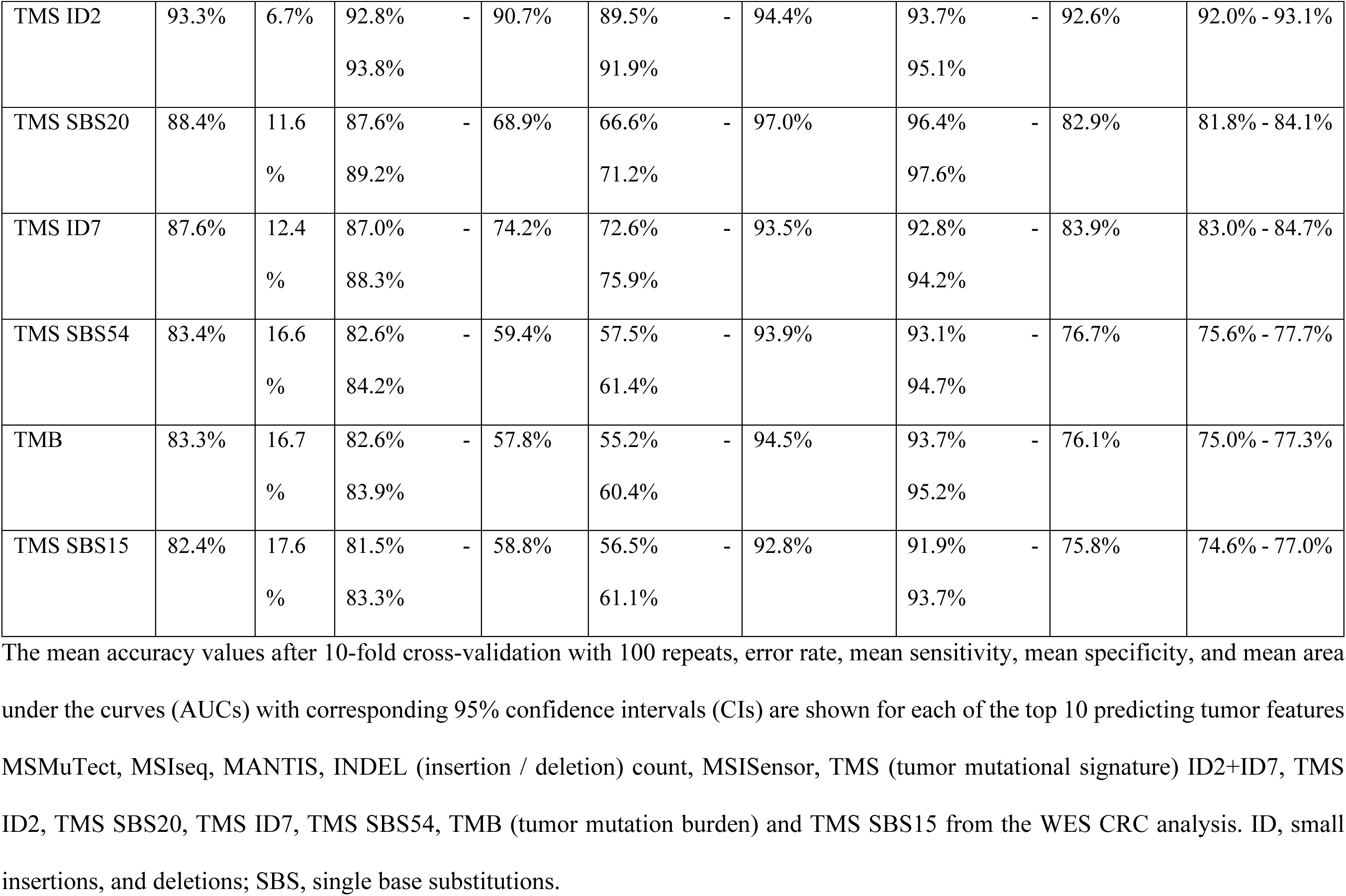
Performance of the top tumor features demonstrating a prediction accuracy >80% ranked by highest mean accuracy from whole-exome sequenced (WES) colorectal cancers (CRCs).

The mean, SD, and range of values for each of these top 10 dMMR predictive features by MMR status and by dMMR subtype for the 300 WES CRCs are shown in **Supplementary Table 4**. For each of these features, the mean values were significantly different between the dMMR and pMMR CRCs (all p<1x10^-12^ from a *two-tailed t-test*), with TMS ID2+ID7 showing the most significant difference (p-value = 7.775x10^-98^), although MSISensor presented with the highest Cohen’s *d* effect size of 4.5, indicating that the means of the pMMR and dMMR groups differed by more than four times the SD (**Supplementary Table 4**). The variation in proportion or counts was larger in the dMMR tumors than in the pMMR tumors for all but one of these top 10 features where TMS ID2+ID7 demonstrated a broad range of values in the pMMR CRCs compared with the dMMR CRCs (**Figure 2, Supplementary Table 4**).

**Figure 2.**
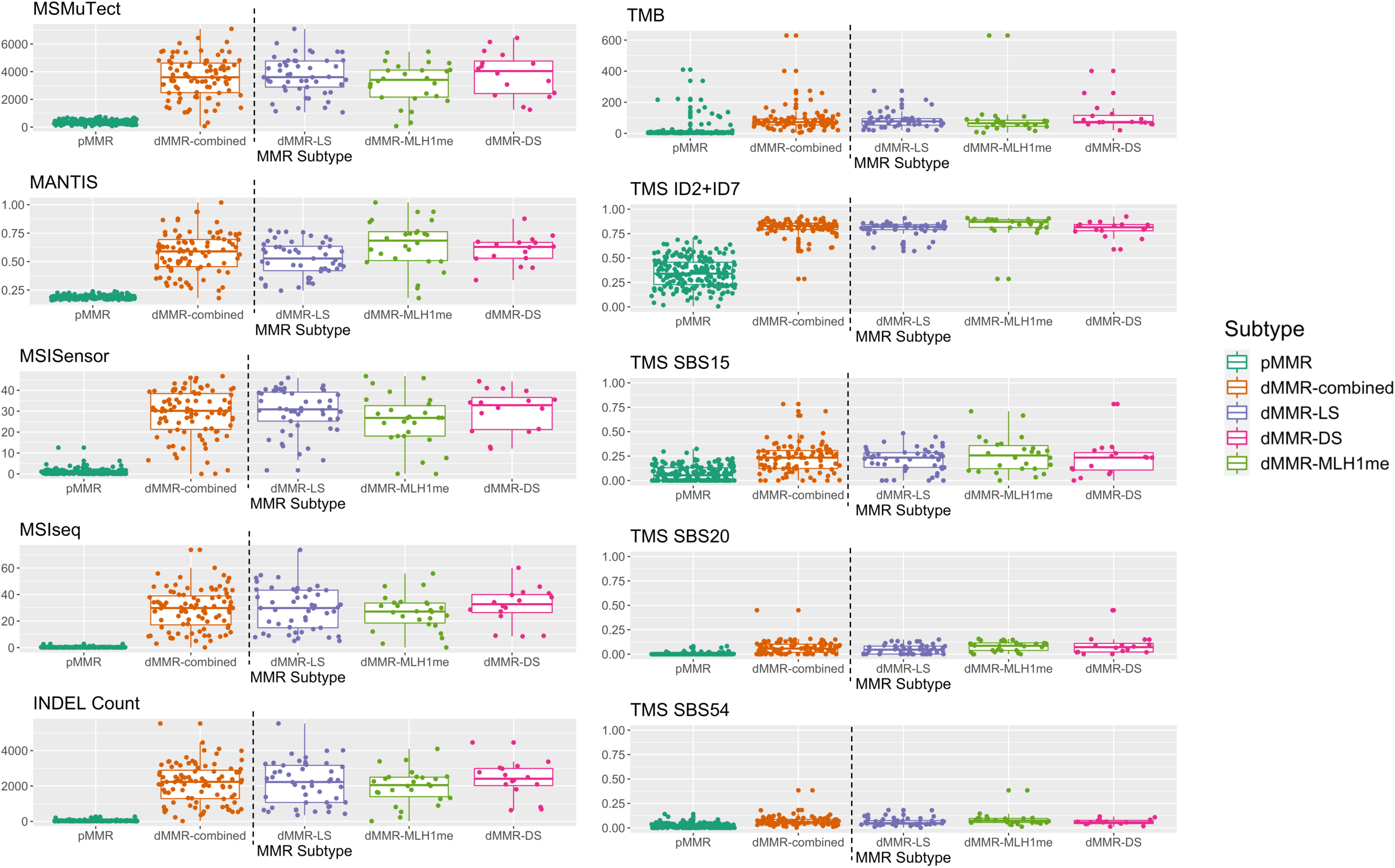
Tumor distribution of the top 10 DNA mismatch repair (MMR) deficient (dMMR) predicting features in the whole exome sequenced (WES) colorectal cancers (CRCs) by MMR subtype. Boxplots showing the distribution of tumors by MMR status (MMR- proficient (pMMR) versus dMMR) as well as stratified by dMMR subtype - dMMR-LS (Lynch syndrome), dMMR-DS (double somatic MMR gene mutations) and dMMR-MLH1me (*MLH1* promoter methylation) for each of the top 10 predicting features MSMuTect, MANTIS, MSISensor, MSIseq, INDEL (insertion / deletion) count, TMB (tumor mutation burden calculated as mutations / mega base), TMS (tumor mutational signature) ID2+ID7, TMS SBS15, TMS SBS20 and TMS SBS54 as determined from the WES CRC analysis. ID, small insertions / deletions; SBS, single base substitution.

The AUCs for the top 10 features when taking all possible thresholds into account are shown in **Supplementary Figure 1.** The MSI prediction tools MSMuTect, MSIseq, and MANTIS as well as INDEL count demonstrated the best AUCs. In addition, we calculated recommended thresholds for each feature for differentiating dMMR from pMMR CRCs using the methodology described in the methods (**Supplementary Table 5**). When applying these thresholds, it was not possible to achieve a complete separation between the dMMR and pMMR tumors for each of the tumor features (**Figure 3**).

**Figure 3.**
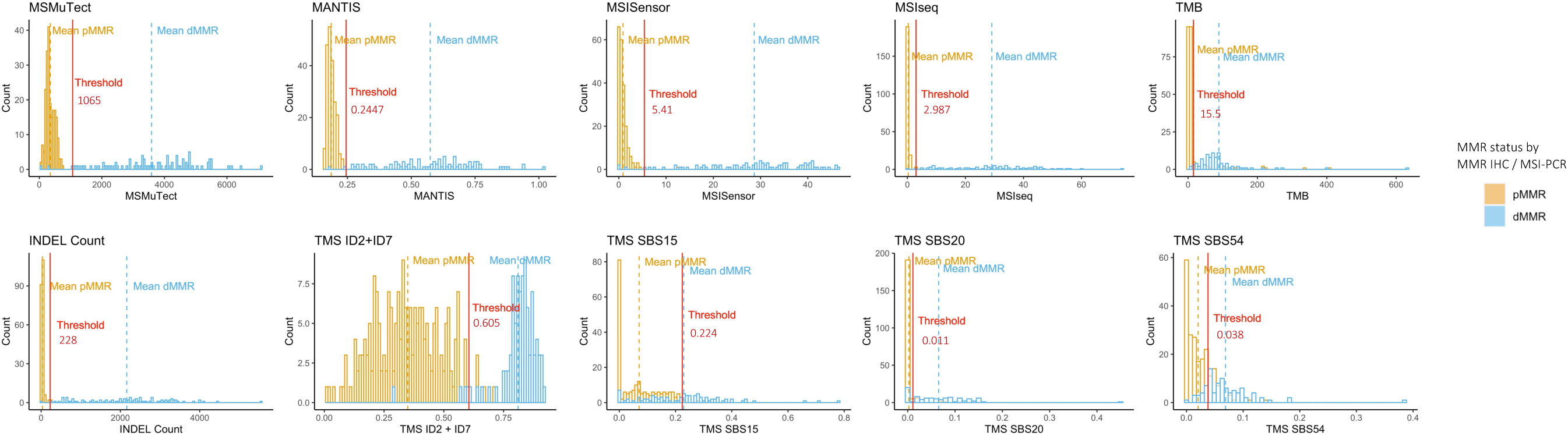
Determination of thresholds for differentiating DNA mismatch repair (MMR) deficient (dMMR) from MMR-proficient (pMMR) colorectal cancers (CRCs) using whole exome sequencing (WES) data for each of the top 10 performing tumor features. Bar graphs presenting the distribution of tumors after applying the recommended thresholds (red line) for each of the top 10 predicting tumor features MSMuTect, MANTIS, MSISensor, MSIseq, INDEL count, TMB, TMS ID2+ID7, TMS SBS15, TMS SBS20 and TMS SBS54 as determined from the WES CRC analysis. Orange coloring indicates pMMR and blue coloring represents dMMR status. ID, small insertions / deletions; SBS, single base substitution.

Investigation of the CRCs misclassified based on the individual tumor feature analysis demonstrated that the misclassification rate (error rate) for the MSI tools was low with MSMuTect (2/300), MANTIS (1/300), MSIseq (1/300) and MSISensor (5/300) calling ≤5 incorrectly out of 300 tumors (≤1.7% error rate). Of the CRCs misclassified by the MSI tools, only two tumors were misclassified by more than one MSI tool, both were dMMR-MLH1me CRCs classified as pMMR. Of note, one of these dMMR-MLH1me CRCs was misclassified as a pMMR tumor by 9 out of the top 10 tumor features. The second misclassified dMMR-MLH1me CRC was classified as pMMR by MSMuTect and MSISensor but classified as dMMR by MSIseq and MANTIS (overall 6/10 features classified this CRC as dMMR). For INDEL count, 3/300 were incorrectly classified, where two pMMR CRCs were classified as dMMR. TMS ID2+ID7 had 10/300 incorrect classifications with seven pMMR tumors incorrectly called as dMMR. The remaining features from the top 10 prediction accuracy list demonstrated the following incorrect classifications: SBS20 (34/300), SBS54 (55/300), SBS15 (44/300) and TMB (19/300) encompassing incorrect calls in both directions (dMMR to pMMR and vice versa).

#### B) Generation of a statistical model by combining tumor features

We assessed whether a combination of features within a statistical model could improve dMMR prediction accuracy. For this, we performed a Lasso penalized logistic regression. Here, after calculating the best lambda value, we found that the combination of TMS ID2+ID7 (coefficient = 5.29), MANTIS (coefficient = 1.70), MSISensor (coefficient = 0.09) with SBS15 (coefficient = 2.25) provided the best prediction accuracy from all possible feature combinations, demonstrating a mean accuracy of 98.3% (95% CI: 0.981-0.986), sensitivity of 0.973 (95% CI: 0.966-0.980) and specificity of 1.000 (95% CI: 1.000-1.000) on the test set.

#### C) Assessing an additive feature combination count for dMMR prediction

Based on the observation that the top performing tumor features from the individual feature analysis did not all misclassify the same CRCs lead us to explore a novel approach of combining tumor features together to increase the overall accuracy i.e., an additive tumor feature combination approach. This approach used a majority count of individual tumor features to overcome the small inaccuracies that each of the top tumor features displayed individually i.e., if one of these top dMMR predictive tumor features misclassified a CRC then the other top dMMR predictive tumor features would correctly classify the same CRC and, thereby, achieve the correct classification overall. Six of the top 10 features from the 10-fold cross-validation analysis demonstrated a mean prediction accuracy of >95% and thus had the least number of incorrect CRC tumor classifications, consisting of MSMuTect, MANTIS, MSIseq, MSISensor, INDEL count, and TMS ID2+ID7. We applied the recommended threshold for determining dMMR status determined previously for each tumor feature (**Figure 3**, **Supplementary Table 5**) to derive a count out of these six selected features, in which each feature is weighted equally. The results show a bimodal distribution across the 300 CRCs (**Figure 4)** where 0/6 to 2/6 features correctly classified all the pMMR CRCs and 4/6 to 6/6 correctly classified all but one of the dMMR tumors with an accuracy of 99.7%. The only exception was the previously mentioned dMMR-MLH1me tumor, which did not meet the recommended thresholds for all six features and thus received a count of 0/6 features suggestive the CRC is pMMR rather than its initial dMMR status.

**Figure 4.**
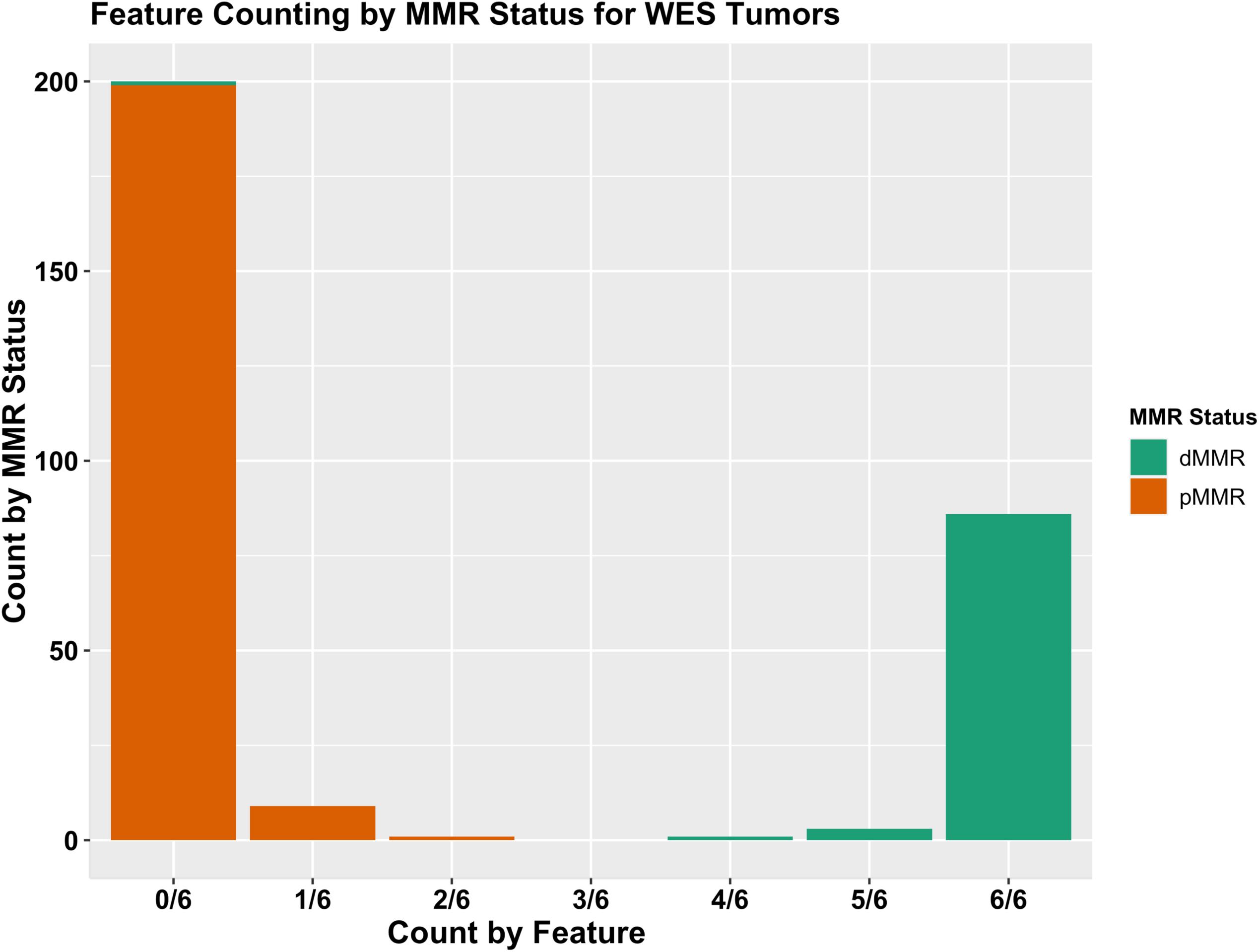
The additive tumor feature combination approach demonstrating the distribution of counts of the top six tumor features by the DNA mismatch repair (MMR) status of the 300 colorectal cancers (CRCs) with whole exome sequencing (WES). Bar graphs presenting the distribution of tumors after applying the additive tumor feature combination approach with the recommended thresholds from the WES CRC analysis using a count of ≥3 out of the top six predictors from the WES CRC analysis, consisting of MSMuTect, MANTIS, MSIseq, MSISensor, INDEL (insertion / deletion) count and TMS (tumor mutational signature) ID2+ID7 (small insertions / deletions) for MMR status calling: MMR-deficient (dMMR) versus MMR-proficient (pMMR).

A summary of the results from the WES CRC analysis for the three approaches is shown in **Table 3** and **Figure 1**.

**Table 3.**
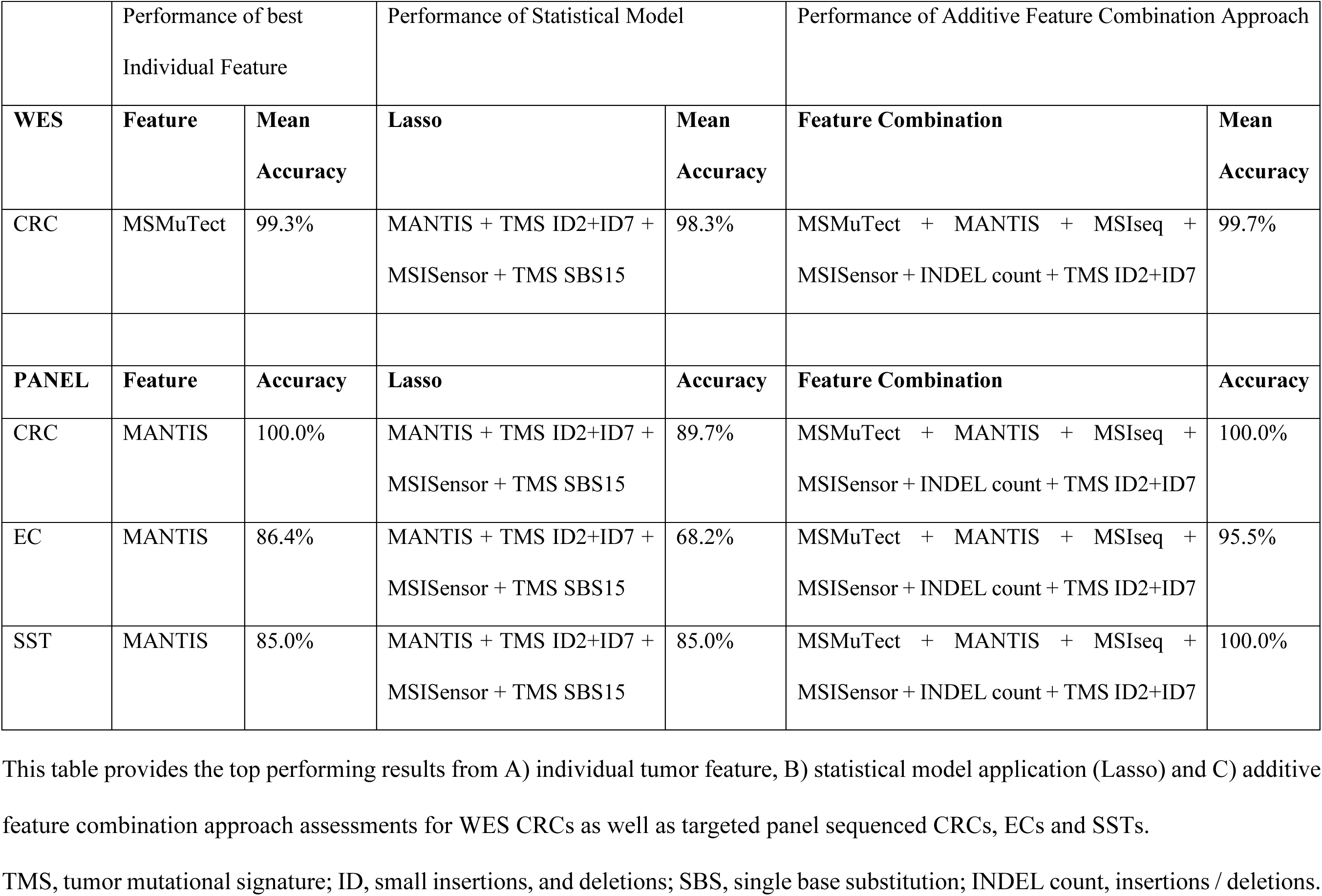
Summary of the best dMMR prediction results by individual tumor feature, Lasso regression model and the additive feature combination approach for the whole-exome sequencing (WES) colorectal cancers (CRCs) and the panel sequenced CRCs, endometrial cancers (ECs) and sebaceous skin tumors (SST).

|  | Performance of best Individual Feature |  | Performance of Statistical Model |  | Performance of Additive Feature Combination Approach |  |
| --- | --- | --- | --- | --- | --- | --- |
|  | Feature | Mean Accuracy | Lasso | Mean Accuracy | Feature Combination | Mean Accuracy |
| WES |  |  |  |  |  |  |
| CRC | MSMuTect | 99.3% | MANTIS + TMS ID2+ID7 + MSISensor + TMS SBS15 | 98.3% | MSMuTect + MANTIS + MSIseq + MSISensor + INDEL count + TMS ID2+ID7 | 99.7% |
| PANEL |  |  |  |  |  |  |
|  | Feature | Accuracy | Lasso | Accuracy | Feature Combination | Accuracy |
| CRC | MANTIS | 100.0% | MANTIS + TMS ID2+ID7 + MSISensor + TMS SBS15 | 89.7% | MSMuTect + MANTIS + MSIseq + MSISensor + INDEL count + TMS ID2+ID7 | 100.0% |
| EC | MANTIS | 86.4% | MANTIS + TMS ID2+ID7 + MSISensor + TMS SBS15 | 68.2% | MSMuTect + MANTIS + MSIseq + MSISensor + INDEL count + TMS ID2+ID7 | 95.5% |
| SST | MANTIS | 85.0% | MANTIS + TMS ID2+ID7 + MSISensor + TMS SBS15 | 85.0% | MSMuTect + MANTIS + MSIseq + MSISensor + INDEL count + TMS ID2+ID7 | 100.0% |

This table provides the top performing results from A) individual tumor feature, B) statistical model application (Lasso) and C) additive feature combination approach assessments for WES CRCs as well as targeted panel sequenced CRCs, ECs and SSTs. TMS, tumor mutational signature; ID, small insertions, and deletions; SBS, single base substitution; INDEL count, insertions / deletions.

### Assessment of individual tumor features, Lasso statistical model and additive feature combination approaches derived from the WES analysis on panel sequenced CRCs, ECs, and SSTs

To determine the generalizability of the findings from the three approaches performed on the WES CRCs, we tested 71 tumors with targeted panel sequencing data to evaluate performance on both a smaller capture and across different tissue types known to have a high prevalence of dMMR.

#### A) Evaluation of the top performing individual features from WES analysis on the panel sequenced CRC, EC, and SST tumors

Out of the top 10 dMMR tumor features from the WES CRC analysis, only four achieved a mean dMMR prediction accuracy of >80% in the panel sequenced CRC tumors (**Table 4**). For EC and SST tumors only one feature (MANTIS) and two features (MANTIS and TMS ID2+ID7), respectively, of the top 10 tumor features achieved a mean dMMR prediction accuracy of >80% (**Table 4**). Across the three tissue types, MANTIS demonstrated the highest mean accuracy, achieving 100% (95% CI: 88.1%-100.0%) accuracy in the panel sequenced CRCs, 86.4% accuracy in ECs (95% CI: 65.1%-97.1%) and 85% accuracy in SSTs (95% CI: 62.1%-96.8%) (**Table 4**). MSMuTect and INDEL count performed poorly in all three panel sequenced tissue types compared with their accuracy in the WES CRCs. MSMuTect and INDEL count are features that provide absolute counts that in our data were two orders of magnitude smaller in the panel sequenced tumors compared with the WES CRCs. The reduction in discriminatory ability is likely related to differences in the size (WES: 67.7 Mb and panel: 2.0 Mb) and location (additional coverage of intronic regions of the MMR genes in the panel capture) of the regions covered by the WES and panel captures resulting in a lower somatic mutation count.

**Table 4.**
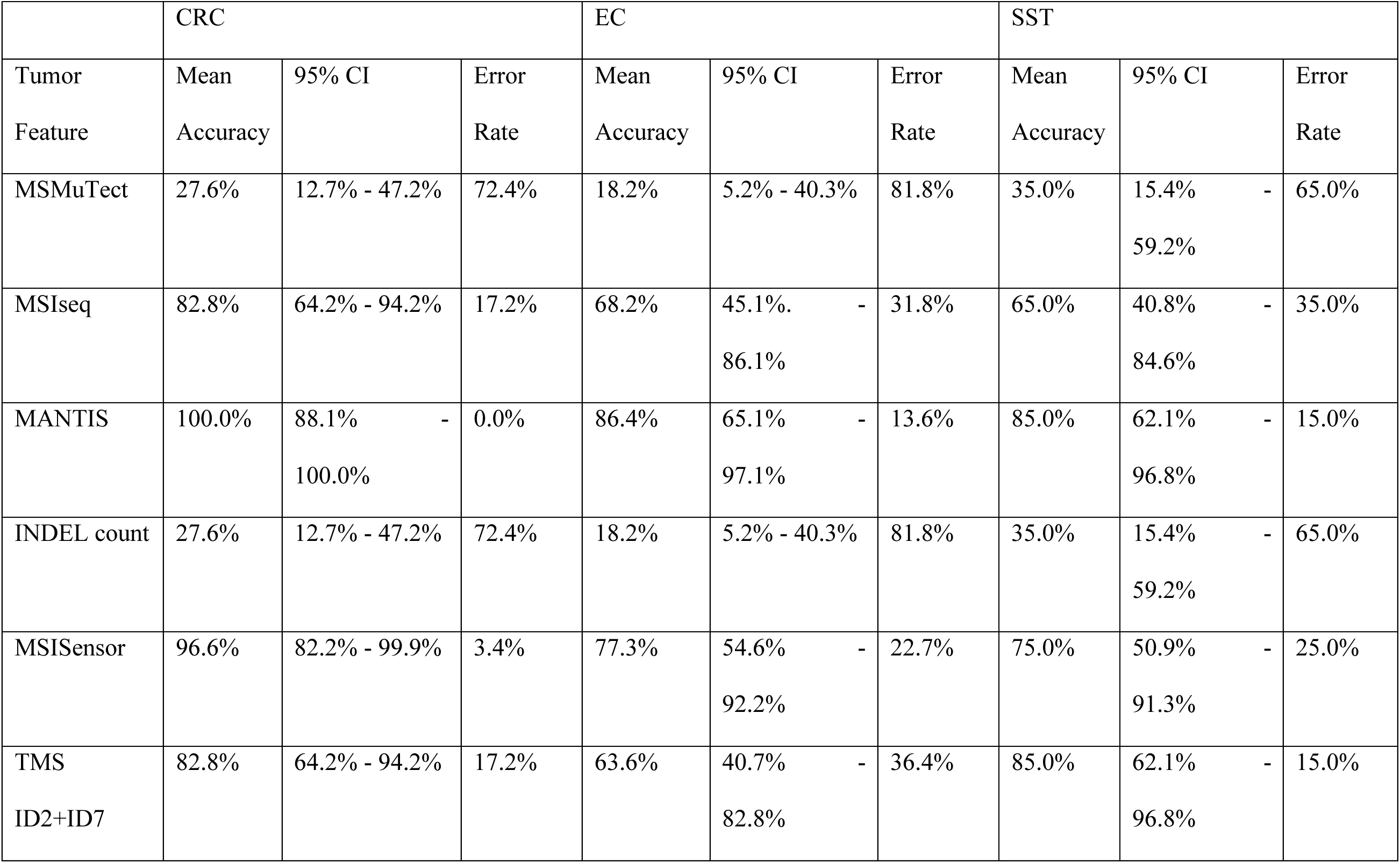
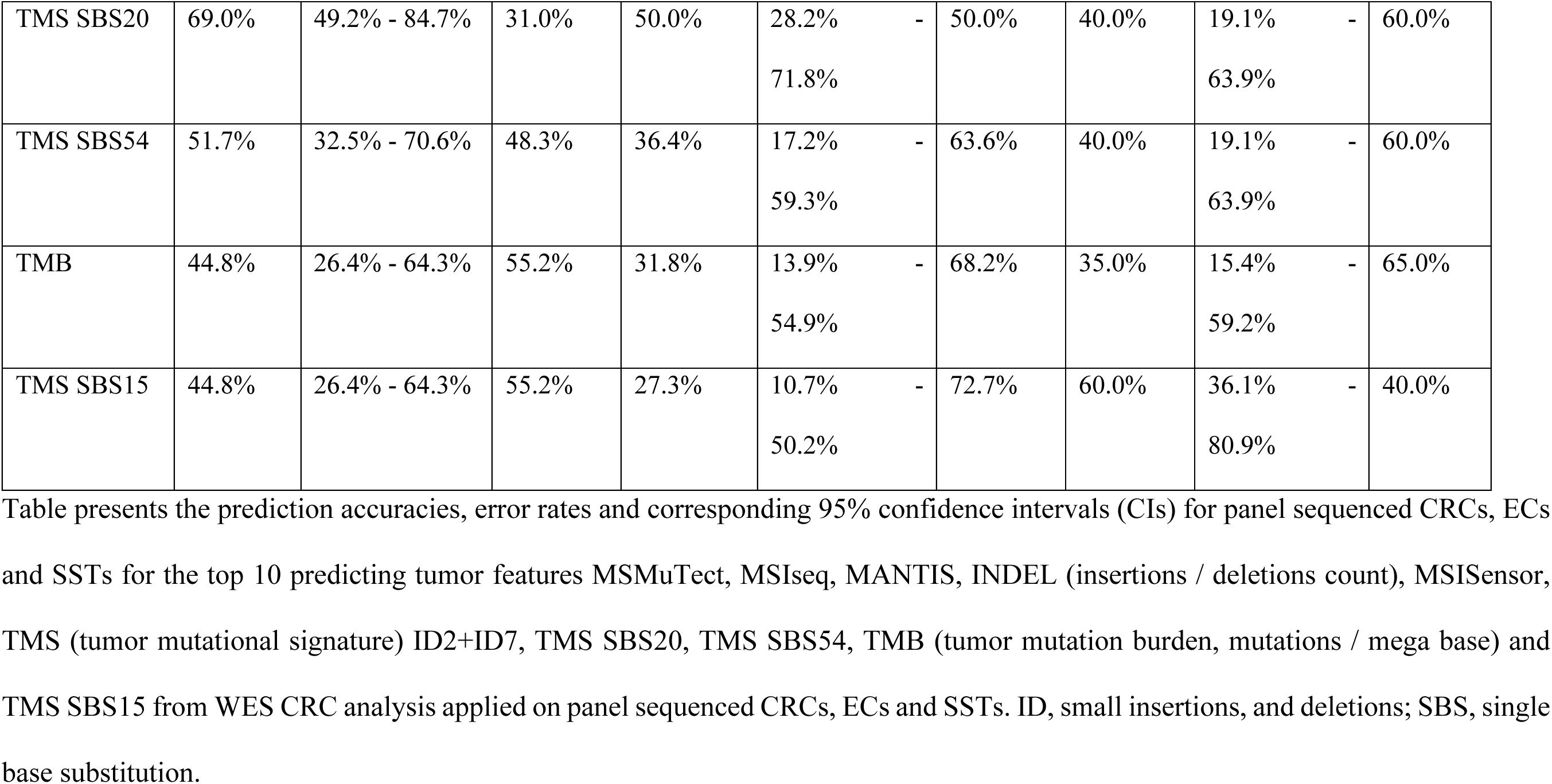
Assessment of top performing tumor features from whole-exome sequenced (WES) colorectal cancers (CRCs) in panel sequenced CRC, endometrial cancer (EC) and sebaceous skin tumor (SST) test sets.

The mean, SD, and range of values for each of these top 10 dMMR predicting features by MMR status and by dMMR subtype for each of CRC, EC and SST tissue types are shown in **Supplementary Tables 6A, 6B, 6C** and in **Supplementary Figure 2, Supplementary Figure 3,** and **Supplementary Figure 4,** respectively. The mean values of each of the top 10 predictors were significantly different between the dMMR and pMMR tumors in all three tissue types except for TMS SBS15 in CRCs, MSISensor in ECs, TMB in ECs and SSTs and, TMS SBS20 and TMS SBS54 in SSTs. MSMuTect consistently had the highest Cohen’s *d* effect size of all top 10 tumor features for each tissue type with the highest effect size observed in CRCs (3.2), indicating the mean of the dMMR and pMMR subgroups for this feature differ by approximately three SDs.

#### B) Evaluation of the Lasso statistical model on the panel sequenced CRC, EC, and SST tumors

From WES analysis, the Lasso statistical model comprised of TMS ID2+ID7, MANTIS, MSISensor and SBS15 achieved a mean prediction accuracy of 98.3%. When this model was applied, with the coefficients determined from the WES analysis, on these three independent panel sequenced tissue types, the prediction accuracies were lower (CRC: 89.7%, EC: 68.2% and SST: 85.0%) (**Table 3**).

#### C) Evaluation of the additive tumor feature combination approach on the panel sequenced CRC, EC, and SST tumors

For each of the top 10 dMMR predictive tumor features we determined the optimal thresholds for the panel sequenced CRCs, ECs, and SSTs (**Supplementary Table 5**) and plotted them by tissue type (CRC - **Supplementary Figure 5**), (EC - **Supplementary Figure 6**), (SST - **Supplementary Figure 7**). The determined thresholds for MANTIS were consistent across both WES and panel captures as well as across tissue types while the calculated thresholds for MSIseq were consistent for CRC across WES and panel captures but different to the thresholds determined for EC and SST. The remaining eight tumor features showed variability in their determined thresholds across both capture type and tissue type (**Supplementary Table 5**). As such, we applied the thresholds determined for each tissue type for the panel sequenced data in the additive feature combination approach below.

The additive feature combination approach incorporates a count of MSMuTect, MANTIS, MSIseq, MSISensor, INDEL count and TMS ID2+ID7 tumor features to classify a tumor as dMMR. The distribution of the counts of these six tumor features determined for each tumor are shown for CRC (**Supplementary Figure 8**), EC (**Supplementary Figure 9**) and SSTs (**Supplementary Figure 10**). For each tissue type, all the dMMR tumors had ≥3/6 tumor features classify them as dMMR, except for a single dMMR-MLH1me EC (1/71, 1.4%) which scored 0/6 and, therefore, was suggestive of pMMR status. This approach achieved accuracy scores of 100%, 95.5% and 100%, for CRC, EC and SST, respectively (**Table 3**).

A summary of the WES CRC and CRC, EC, and SST panel sequencing results for all three approaches is provided in **Table 3**.

## Discussion

In this study, we compared tumor features calculated from next generation sequencing data for their accuracy in predicting dMMR status in 300 CRCs, 91 of which were dMMR determined by immunohistochemistry or MSI-PCR and with an established sporadic or inherited etiology for their dMMR status. Ten features achieved >80% dMMR prediction accuracy from the WES CRC tumors, with the highest accuracy predictors being the MSI tools MSMuTect, MSIseq and MANTIS, all of which achieved ≥99% accuracy. The combination of TMS ID2+ID7 achieved the highest mean accuracy for dMMR prediction out of the 97 TMS features assessed. When applied to the targeted multi-gene panel setting, the performance of these 10 features was reduced not only in CRC but also for the EC and SST tumors. In addition, we investigated two approaches that combined these top 10 performing tumor features to improve the overall prediction accuracy. The Lasso generated model achieved 98.3% accuracy in WES CRCs although the performance of the model was reduced in the panel sequenced CRC, EC, and SST tumors. For both the WES CRCs and panel sequencing across tissue types, the additive tumor feature combination approach, where having ≥3 of the top 6 tumor features classify a tumor as dMMR, achieved the highest prediction accuracies of the three approaches tested.

To date, multiple tools to detect MSI from NGS data have been developed^71^. NGS based MSI tool development has been constantly evolving since the introduction of MSISensor^28^ and mSINGS^72^, which were followed by MSIseq^29^, MANTIS^30^ and MSMuTect^31^. However, to the best of our knowledge, neither a comparison of more than three MSI detection tools on the same tumor sample nor the effectiveness of these MSI tools specifically on SST tumors has been performed to date. Previously, MANTIS has been compared to MSISensor with the former showing superior sensitivity (97.18% vs. 96.48%) and specificity (99.68% vs. 98.73%)^30^. This was supported by our findings, and we additionally showed that across the WES and panel tested CRCs, MANTIS provided the highest dMMR prediction accuracy and was shown to be the top performing feature in the EC and SST tumors as well. Recently, the United States Food and Drug Administration (FDA) approved MSISensor for detecting MSI in metastatic CRCs for selecting patients for immune checkpoint inhibition therapy^73^. In our study, MSISensor had the lowest accuracy (97.7%) in WES CRCs of the four MSI tools tested, incorrectly classifying 5/300 CRCs. Seeking FDA approval for other MSI tools in addition to MSISensor is warranted based on our findings.

MSMuTect has been trained on 20 different tissue types using WES data and, therefore, it was not surprising it had the highest mean accuracy of the top performing tumor features in our WES CRC analysis. MSMuTect has been designed to accurately detect somatic MSI indels using a count of indels from the captured sequencing region^31^. Thus, the MSI indel count from WES data (67.7 Mb) could be up to ∼34x larger than that from panel data (2.0 Mb), which likely explains the poor performance of this tool observed in our panel sequencing data test sets. When we adjusted the MSMuTect threshold for calling dMMR for panel data, MSMuTect showed improved discrimination of dMMR from pMMR tumors. This increase in prediction accuracy was also observed for the INDEL count where adjusting the threshold for panel data improved the overall performance. Adjusting the threshold for panel sequencing data enabled the inclusion of MSMuTect and INDEL count as two of the six tumor features in our additive feature combination approach that ultimately performed well on panel sequenced tumors. Tumor features that calculate a percentage rather than raw counts such as MANTIS, MSISensor, SBS TMS and ID TMS are more adaptable to changes in capture size. For example, our results showed that the calculated thresholds for differentiating dMMR from pMMR for MANTIS were consistent across both WES and panel captures as well as across tissue types. Therefore, we recommend training features that incorporate a count of genomic variants, such as INDELs, SNVs and MSMuTect on the capture size to improve dMMR prediction accuracy.

While three ID TMS (ID1, ID2 and ID7) are reported to be associated with dMMR^33^, our results showed that the combination of ID2 and ID7 TMS achieved the highest dMMR prediction accuracy of any of the TMS features in WES CRC tumors, outperforming ID2 or ID7 alone. Of the seven SBS TMS that are associated with dMMR (SBS6, SBS14, SBS16, SBS20, SBS21, SBS26 and SBS44)^33^, only two, TMS SBS15 and TMS SBS20, showed >80% dMMR prediction accuracy in WES CRC tumors, but were shown to be poor predictors in the panel sequenced tumors. Interestingly, TMS SBS54 was one of the top 10 dMMR predictors from the WES CRC analysis, although currently its proposed etiology in COSMIC is related to a “possible sequencing artefact and/or a possible contamination with germline variants”^33^. Another study has shown that SBS15, SBS20 and SBS54 are observed in CRCs with a high immune cytolytic activity (CYT) compared with CYT-low CRCs^74^. CYT-high CRCs have been shown to correlate with an increased somatic mutation load and high levels of MSI^75^, this may explain the observation of TMS SBS15, TMS SBS20 and TMS SBS54 demonstrating >80% dMMR prediction accuracy in our WES CRC analysis.

The combination of tumor features via the Lasso regression model achieved similar mean accuracy as the four MSI tools individually in the WES CRC analysis. The Lasso calculated final model that best distinguished dMMR from pMMR tumors in the WES CRC cohort consisted of TMS ID2+ID7, MANTIS, MSISensor and TMS SBS15. The statistical approach used to determine the final model assigns a ‘weight’ (coefficient value) or confidence of how well each feature detects dMMR. As per generalized linear modelling methodology, the weight of any given feature is reduced as the model incorporates additional features. Hence, with MANTIS being one of the best predictors, its weighting was reduced when other features were added to the final model. This resulted in the Lasso model prediction accuracy being lower than MANTIS alone. Of note, since most of the approaches taken (i.e., assessing features individually or in combination) already achieved a very high prediction accuracy of ∼99%, alternate modelling approaches such as Random Forest would not result in a significant improvement in dMMR prediction accuracy.

Strengths of our study were a large sample of tumors including dMMR tumors with confirmed sporadic or inherited etiology concordant with MMR IHC and MSI-PCR results for both the WES and panel sequenced datasets. Tumor MMR status combined with identified etiology provided a more reliable reference group of CRCs than would a group based on MMR IHC test results without etiological confirmation given the known challenges that can lead to false positive and negative MMR IHC results^16^. We assessed many tumor features that can be readily derived from NGS data ensuring that our findings have potential to be easily implemented in clinical diagnostics. We applied our findings from WES to panel data to determine the generalizability of our findings to smaller panel captures such as those that are currently used in clinical diagnostics. We showed the applicability of our findings on tissue types that display a high proportion of dMMR phenotype. Our dMMR tumor samples included those with the frequent pattern of MMR IHC namely MLH1/PMS2 loss and MSH2/MSH6 loss but also tumors with solitary MSH6 loss or solitary PMS2 loss, ensuring we covered the spectrum of dMMR tissue types which is particularly relevant given the identified challenges associated with interpretation of solitary MSH6 loss^76^.

There were several limitations of our study including testing of only three tissue types. Testing of these tumor features and approaches in other tissue types such as stomach cancer, which also has a high prevalence of dMMR overall and dMMR related to Lynch syndrome, would determine the suitability of these tumor features for inclusion in an additive feature combination approach in a pan-cancer setting. In addition, the sample size for the panel sequenced tumors was limited for all three tissue types, however, there was a high proportion of dMMR in the tumors tested (72.4% for CRC, 81.8% for EC and 65.0% for SST). No tumor feature or approach achieved 100% accuracy in the CRC WES analysis. This was largely related to a single tumor (dMMR-MLH1me) from the WES CRC analysis that was called incorrectly by 9/10 top individual tumor features suggesting the CRC was pMMR. Therefore, we repeated the *MLH1* methylation testing for this tumor using both MethyLight and MS-HRM assays. Both assays found no evidence of *MLH1* methylation in the tumor. These new *MLH1* methylation results and the pMMR classification from our analysis suggest the initial dMMR classification was a false positive. If this CRC would initially have been categorized as a pMMR tumor, then MANTIS and MSIseq would have achieved 100% accuracy in the WES CRC analysis. Furthermore, the identification of an initial tumor misclassification provides strong support for evaluating multiple dMMR prediction tumor features and highlights the advantage of combining these features through an additive feature combination approach.

## Conclusion

Our findings provide an important comparison of tumor features for dMMR prediction, highlighting performance differences between capture size and tissue types. Our results demonstrate the high accuracy of multiple individual tumor features including the MSI calling tools MSMuTect, MSIseq, MANTIS and MSISensor, as well as INDEL count and the combination of TMS ID2+ID7 for predicting dMMR status using WES CRCs. Moreover, our findings highlight the benefit of combining these six tumor features in a simple additive feature combination approach to improve dMMR prediction accuracy, particularly in targeted panel sequencing data from CRC, EC, or SST tumors. With the reported inaccuracies of MMR IHC and the increasing application of clinical NGS testing of tumor tissue, accurately deriving dMMR status from this NGS data will have important implications for diagnostics and targeted therapy and likely improve patient outcomes and cancer prevention.

## Supporting information

Conflict of interest disclosure

STARD checklist

## Data Availability

The datasets generated during and/or analyzed during the current study are available from the corresponding author on reasonable request.

## Acknowledgements

We thank members of the Colorectal Oncogenomics Group and members from the Genomic Medicine and Family Cancer Clinic for their support of this manuscript. We thank the participants and staff from the Australasian and Ontario Colorectal Cancer Family Registries (ACCFR/OFCCR) and the ANGELS, Muir-Torre and WEHI studies. We especially thank Maggie Angelakos, Samantha Fox, Allyson Templeton for supporting this study. We thank the Australian Genome Research Facility for their collaboration on this project. We thank A/Prof Sue Finch of the Melbourne Statistical Consulting Platform and Statistical Consulting Centre at the University of Melbourne for guidance with the statistical aspects of this study.

## Declared conflicts of interest

The authors have no conflicts of interest to declare.

