## Supplementary Figures for "Evaluating multiple next-generation sequencing derived tumor features to accurately predict DNA mismatch repair status"

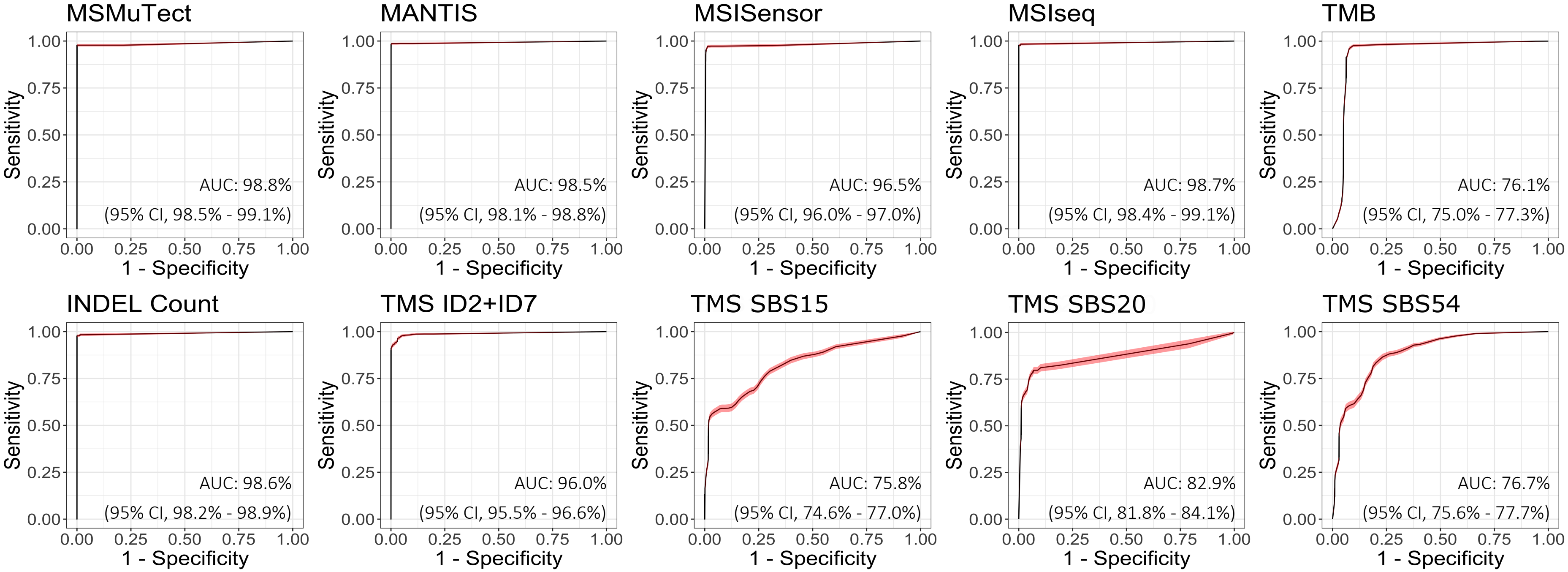


**Supplementary Figure 1.** The cross validated area under the curves (AUCs) for the top 10 performing tumor features to predict the DNA mismatch repair (MMR) status in the whole exome sequenced (WES) colorectal cancer (CRC) dataset. Receiving operator curves (ROCs) presenting the AUCs for the top 10 performing tumor features MSMuTect, MANTIS, MSISensor, MSIseq, INDEL (insertions / deletions) count, TMB (tumor mutation burden calculated as mutations per mega base), TMS (tumor mutational signature) ID2+ID7, TMS SBS15, TMS SBS20 and TMS SBS54 as determined from the WES CRC analysis. The 95% confidence intervals (CIs) for each feature are shown in red highlighting. ID, small insertions / deletions; SBS, single base substitution.

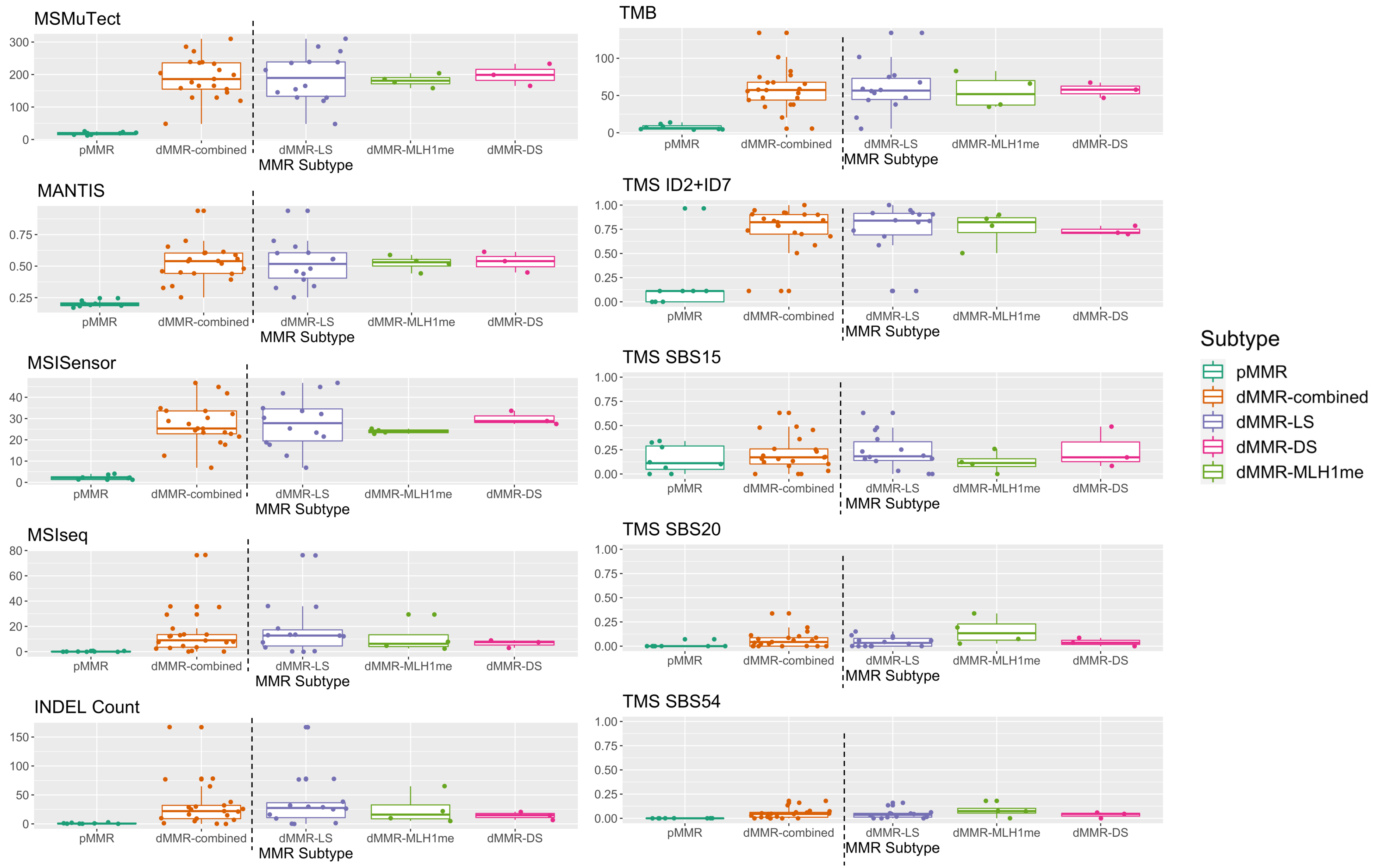
MSMuTect
MANTIS
MSISensor
MSIseq
INDEL Count
TMB
TMS ID2+ID7
TMS SBS15
TMS SBS20
TMS SBS54
pMMR
dMMR-combined
dMMR-LS
dMMR-MLH1me
dMMR-DS
MMR Subtype
Subtype

**Supplementary Figure 2.** Tumor distribution of the top 10 best predicting features from the whole exome sequencing (WES) analysis on the colorectal cancer (CRC) test set. Boxplots showing the distribution of tumors by DNA mismatch repair (MMR) proficient (pMMR) / MMR-deficient (dMMR) subgroups as well as stratified by dMMR subtype - dMMR-LS (Lynch syndrome), dMMR-DS (double somatic MMR gene mutation) and dMMR-MLH1me (*MLH1* promoter methylation) for each of the top 10 predicting features MSMuTect, MANTIS, MSISensor, MSIseq, INDEL (insertions / deletions) count, TMB (tumor mutation burden calculated as mutations per megabase), TMS (tumor mutational signature) ID2+ID7, TMS SBS15, TMS SBS20 and TMS SBS54 from the WES CRC analysis applied on targeted panel sequenced CRCs. ID, small insertions / deletions; SBS, single base substitution.

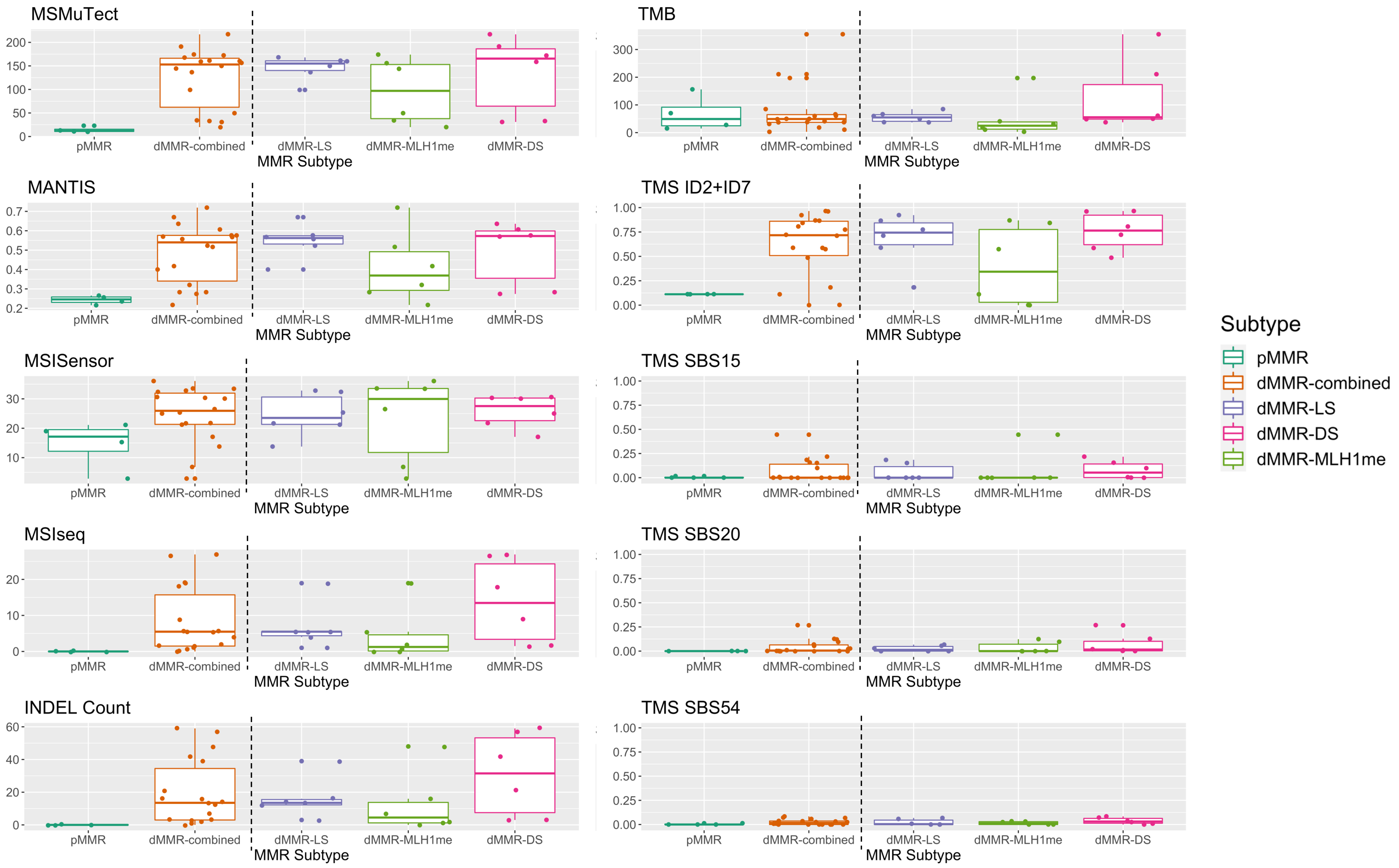

MSMuTect
MANTIS
MSISensor
MSIseq
INDEL Count
TMB
TMS ID2+ID7
TMS SBS15
TMS SBS20
TMS SBS54
pMMR
dMMR-combined
dMMR-LS
dMMR-MLH1me
dMMR-DS
MMR Subtype
Subtype
pMMR
dMMR-combined
dMMR-LS
dMMR-DS
dMMR-MLH1me

**Supplementary Figure 3.** Tumor distribution of the top 10 best predicting features from the whole exome sequencing (WES) analysis on the endometrial cancer (EC) test set. Boxplots showing the distribution of tumors by DNA mismatch repair (MMR) proficient (pMMR) / MMR-deficient (dMMR) subgroups as well as separated by dMMR-LS (Lynch syndrome), dMMR-DS (double somatic MMR gene mutations) and dMMR-MLH1me (*MLH1* promoter methylation) individual subtypes for the top 10 predicting features MSMuTect, MANTIS, MSISensor, MSIseq, INDEL (insertions / deletions) count, TMB (tumor mutation burden calculated as mutations per mega base), TMS (tumor mutational signature) ID2+ID7, TMS SBS15, TMS SBS20 and TMS SBS54 from the WES CRC analysis applied on targeted panel sequenced ECs. ID, small insertions / deletions; SBS, single base substitution.

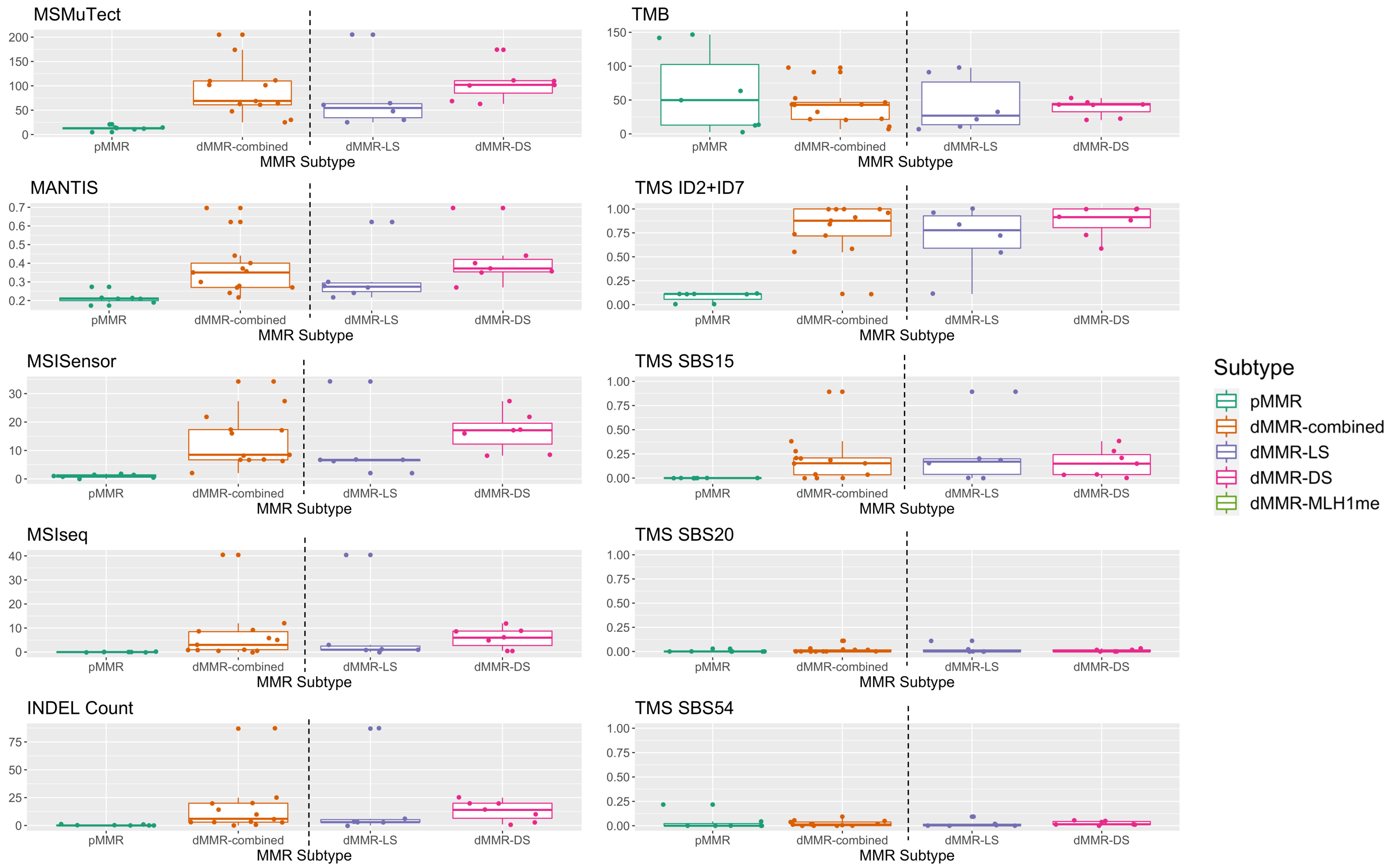

MSMuTect
TMB
MANTIS
TMS ID2+ID7
MSISensor
TMS SBS15
MSIseq
TMS SBS20
INDEL Count
TMS SBS54
pMMR
dMMR-combined
dMMR-LS
dMMR-DS
MMR Subtype
Subtype
pMMR
dMMR-combined
dMMR-LS
dMMR-DS
dMMR-MLH1me

**Supplementary Figure 4.** Tumor distribution of the top 10 best predicting features from the whole exome sequencing (WES) analysis on the sebaceous skin tumor (SST) test set. Boxplots showing the distribution of tumors by DNA mismatch repair (MMR) proficient (pMMR) / MMR-deficient (dMMR) subgroups as well as separated by dMMR-LS (Lynch syndrome), dMMR-DS (double somatic MMR gene mutation) and dMMR-MLH1me (*MLH1* promoter methylation) individual subtypes for the top 10 predicting features MSMuTect, MANTIS, MSISensor, MSIseq, INDEL (insertions / deletions) count, TMB (tumor mutation burden calculated as mutations per mega base), TMS (tumor mutational signature) ID2+ID7, TMS SBS15, TMS SBS20 and TMS SBS54 from the WES CRC analysis applied on targeted panel sequenced SSTs. ID, small insertions / deletions; SBS, single base substitution.

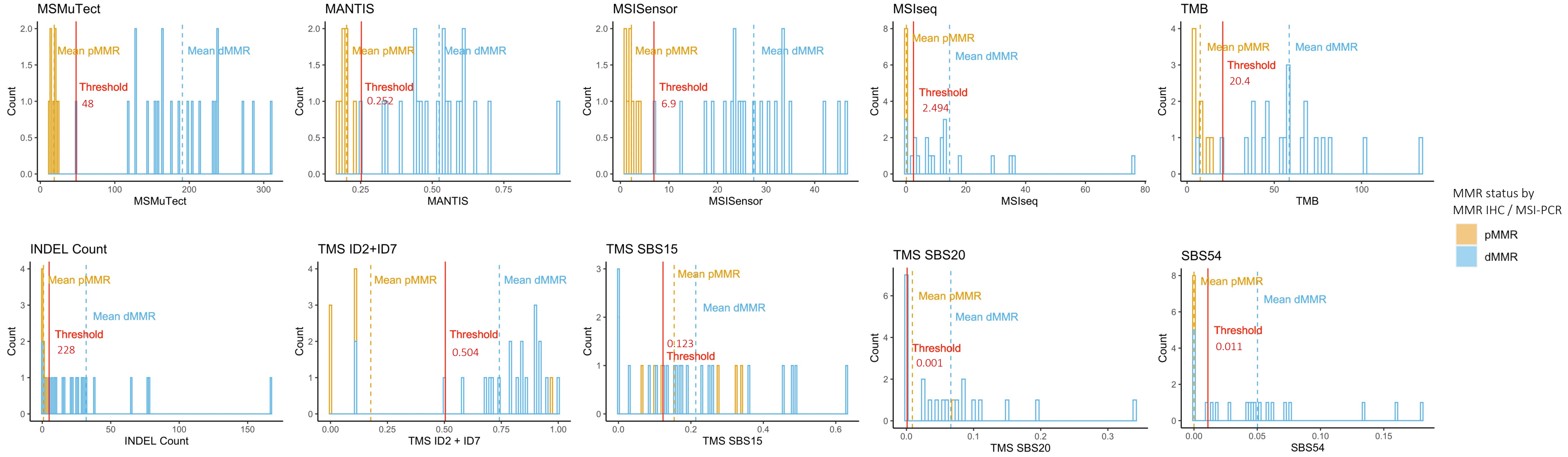


**Supplementary Figure 5.** Determination of the thresholds for the top 10 performing features for panel colorectal cancers (CRCs). Bar graphs presenting the distribution of tumors after applying the recommended thresholds for each of the top 10 predicting tumor features MSMuTect, MANTIS, MSISensor, MSIseq, INDEL (insertions / deletions) count, TMB (tumor mutation burden calculated as mutations per mega base), TMS (tumor mutational signature) ID2+ID7, TMS SBS15, TMS SBS20 and TMS SBS54 for targeted panel sequenced CRCs. Orange coloring indicates DNA mismatch repair (MMR) proficient (pMMR) and blue coloring represents MMR-deficient (dMMR) status. ID, small insertions / deletions; SBS, single base substitution.

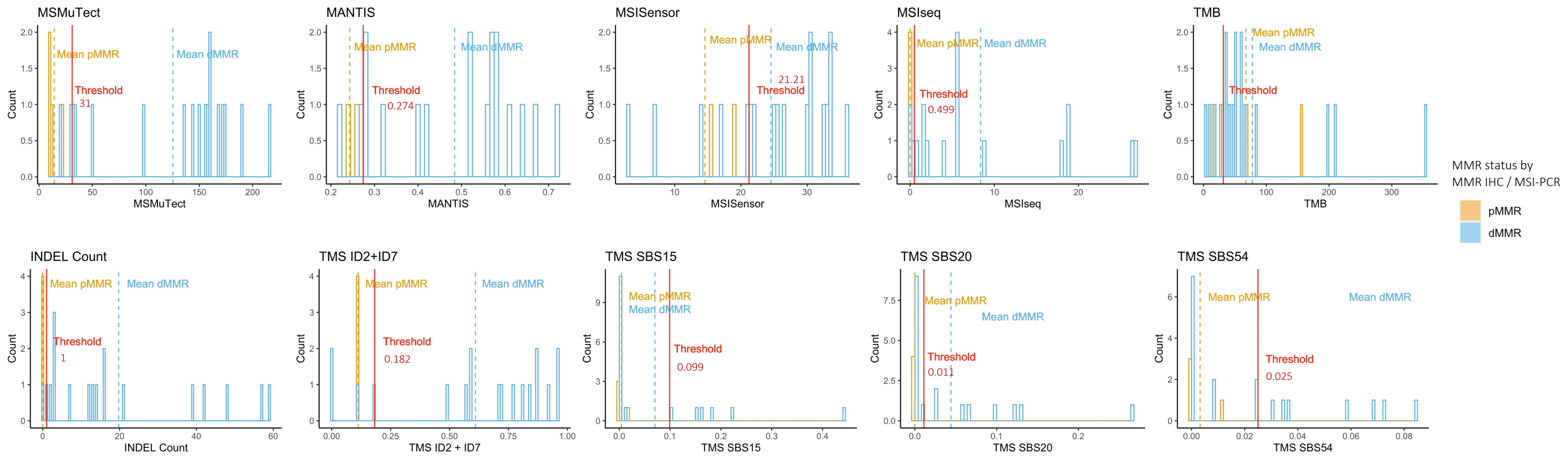


**Supplementary Figure 6.** Determination of the thresholds for the top 10 performing features for panel endometrial cancers (ECs). Bar graphs presenting the distribution of tumors after applying the recommended thresholds for each of the top 10 predicting tumor features MSMuTect, MANTIS, MSISensor, MSIseq, INDEL (insertions / deletions) count, TMB (tumor mutation burden calculated as mutations per megabase), TMS (tumor mutational signature) ID2+ID7, TMS SBS15, TMS SBS20 and TMS SBS54 for targeted panel sequenced ECs. Orange coloring indicates DNA mismatch repair (MMR) proficient (pMMR) and blue coloring represents MMR-deficient (dMMR) status.ID, small insertions / deletions; SBS, single base substitution.

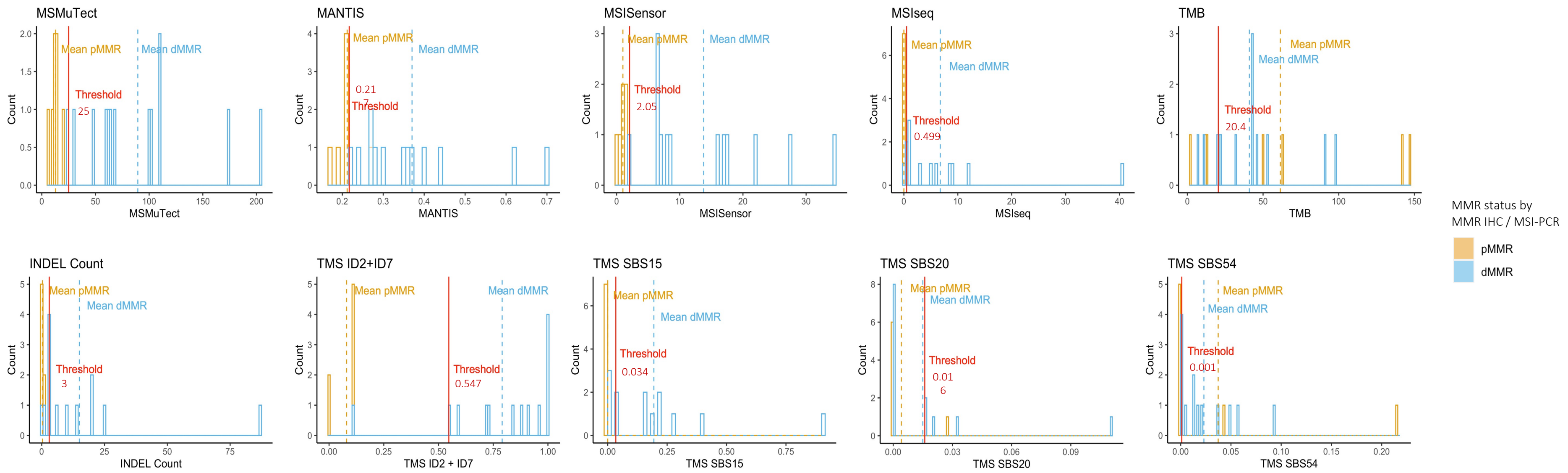


**Supplementary Figure 7.** Determination of the thresholds for the top 10 performing features for panel sebaceous skin tumors (SSTs). Bar graphs presenting the distribution of tumors after applying the recommended thresholds for each of the top 10 predicting tumor features MSMuTect, MANTIS, MSISensor, MSIseq, INDEL (insertions / deletions) count, TMB (tumor mutation burden calculated as mutations per megabase), TMS (tumor mutational signature) ID2+ID7, TMS SBS15, TMS SBS20 and TMS SBS54 for targeted panel sequenced SSTs. Orange coloring indicates DNA mismatch repair (MMR) proficient (pMMR) and blue coloring represents MMR-deficient (dMMR) status.ID, small insertions / deletions; SBS, single base substitution.

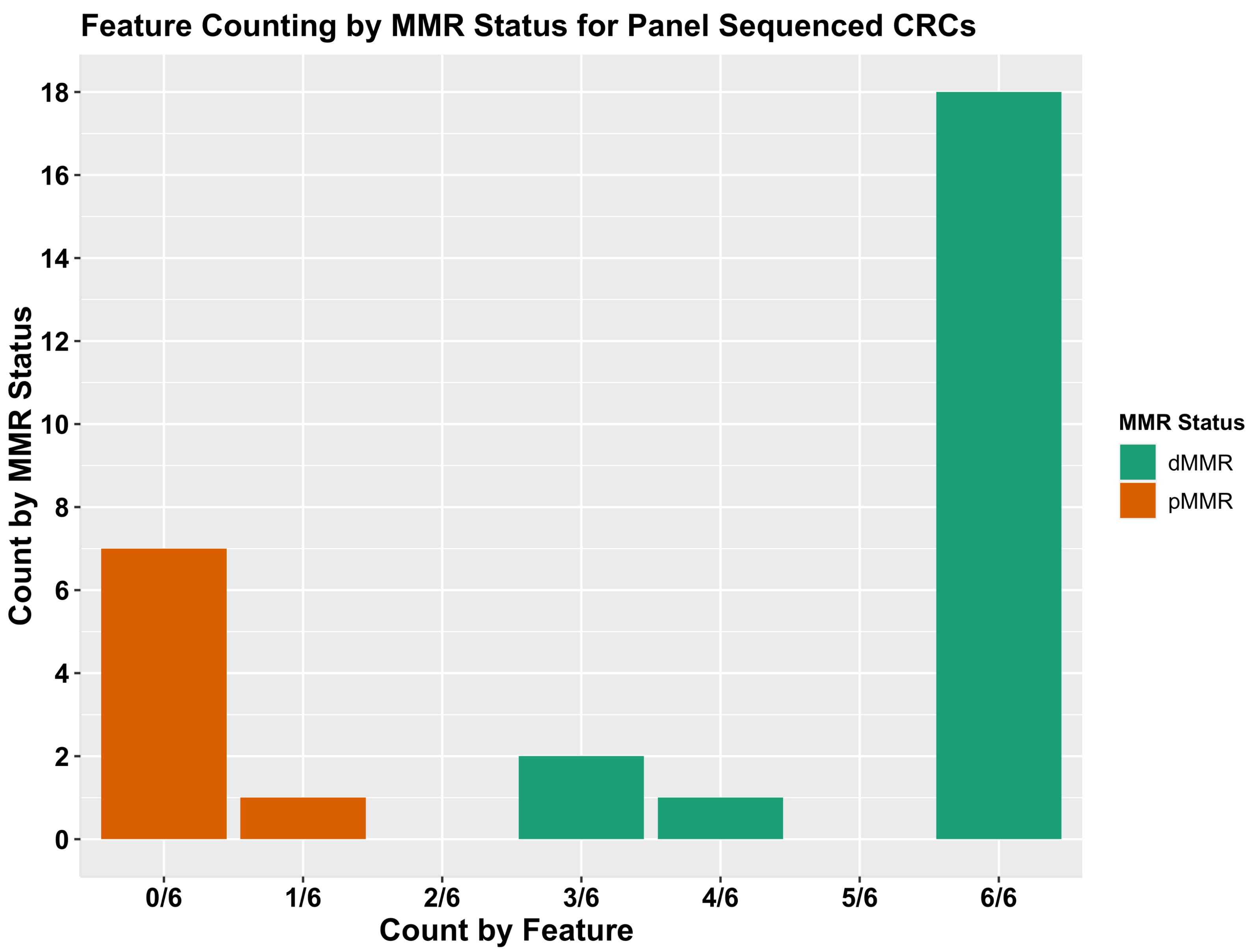

Feature Counting by MMR Status for Panel Sequenced CRCs
Count by MMR Status
0
2
4
6
8
10
12
14
16
18
0/6
1/6
2/6
3/6
4/6
5/6
6/6
Count by Feature
MMR Status
dMMR
pMMR

**Supplementary Figure 8.** The additive tumor feature combination approach demonstrating the distribution of counts of the top six tumor features by the DNA mismatch repair (MMR) status of the 29 panel sequenced colorectal cancers (CRCs). Bar graphs presenting the distribution of tumors after applying the additive tumor feature combination approach with the recommended thresholds for targeted panel sequenced CRCs using a count of ≥3 out of the top six predictors with >95% accuracy consisting of MSMuTect, MANTIS, MSIseq, MSISensor, INDEL (insertions / deletions) count and TMS (tumor mutational signature) ID2+ID7 (small insertions / deletions) for MMR status calling: MMR-deficient (dMMR) versus (MMR-proficient) pMMR.

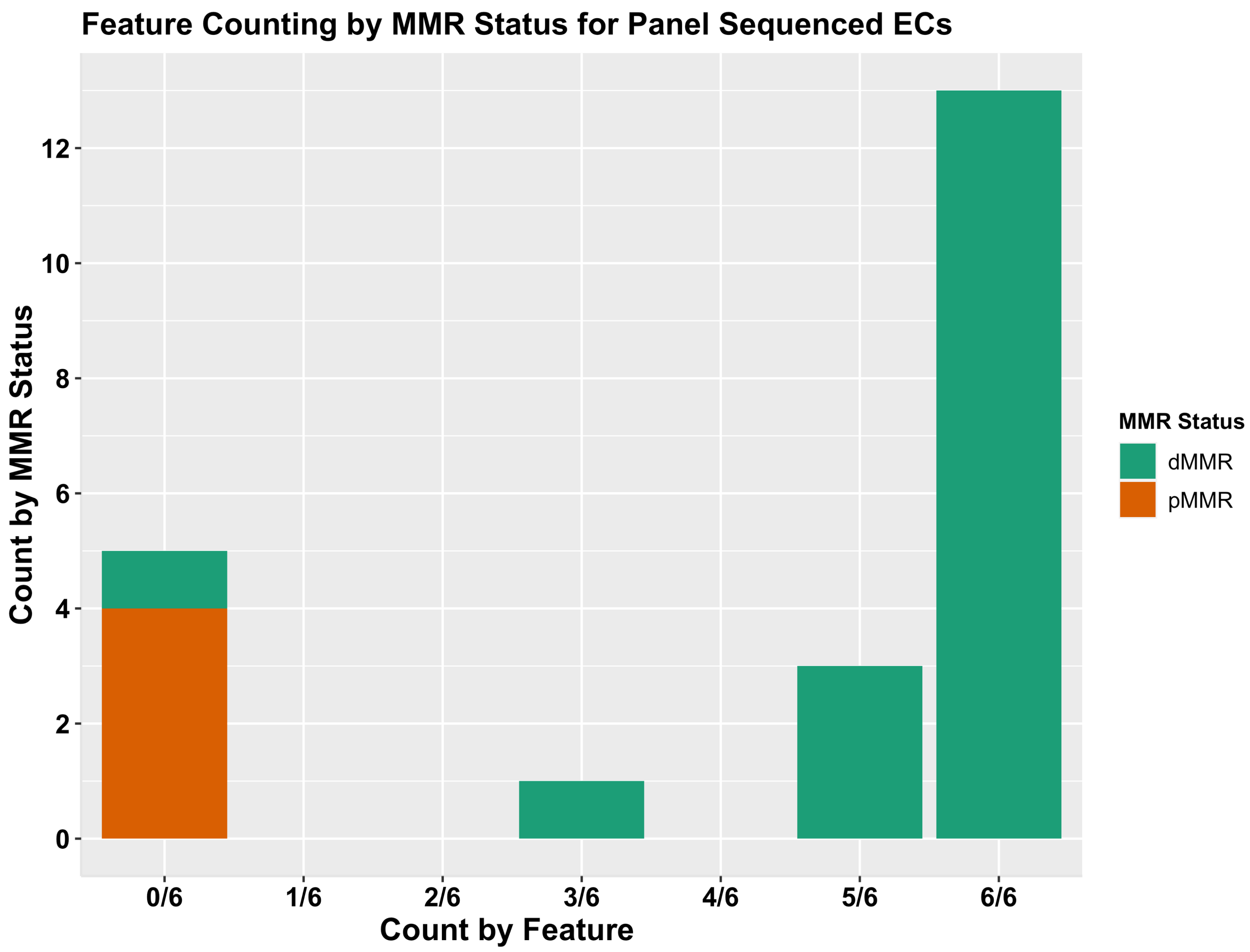
Feature Counting by MMR Status for Panel Sequenced ECs
Count by MMR Status
0
2
4
6
8
10
12
0/6
1/6
2/6
3/6
4/6
5/6
6/6
Count by Feature
MMR Status
dMMR
pMMR

**Supplementary Figure 9.** The additive tumor feature combination approach demonstrating the distribution of counts of the top six tumor features by the DNA mismatch repair (MMR) status of the 22 panel sequenced endometrial cancers (ECs). Bar graphs presenting the distribution of tumors after applying the additive tumor feature combination approach with the recommended thresholds for targeted panel sequenced ECs using a count of ≥3 out of the top six predictors with >95% accuracy consisting of MSMuTect, MANTIS, MSIseq, MSISensor, INDEL (insertions / deletions) count and TMS (tumor mutational signature) ID2+ID7 (small insertions / deletions) for MMR status calling: MMR-deficient (dMMR) versus (MMR-proficient) pMMR.

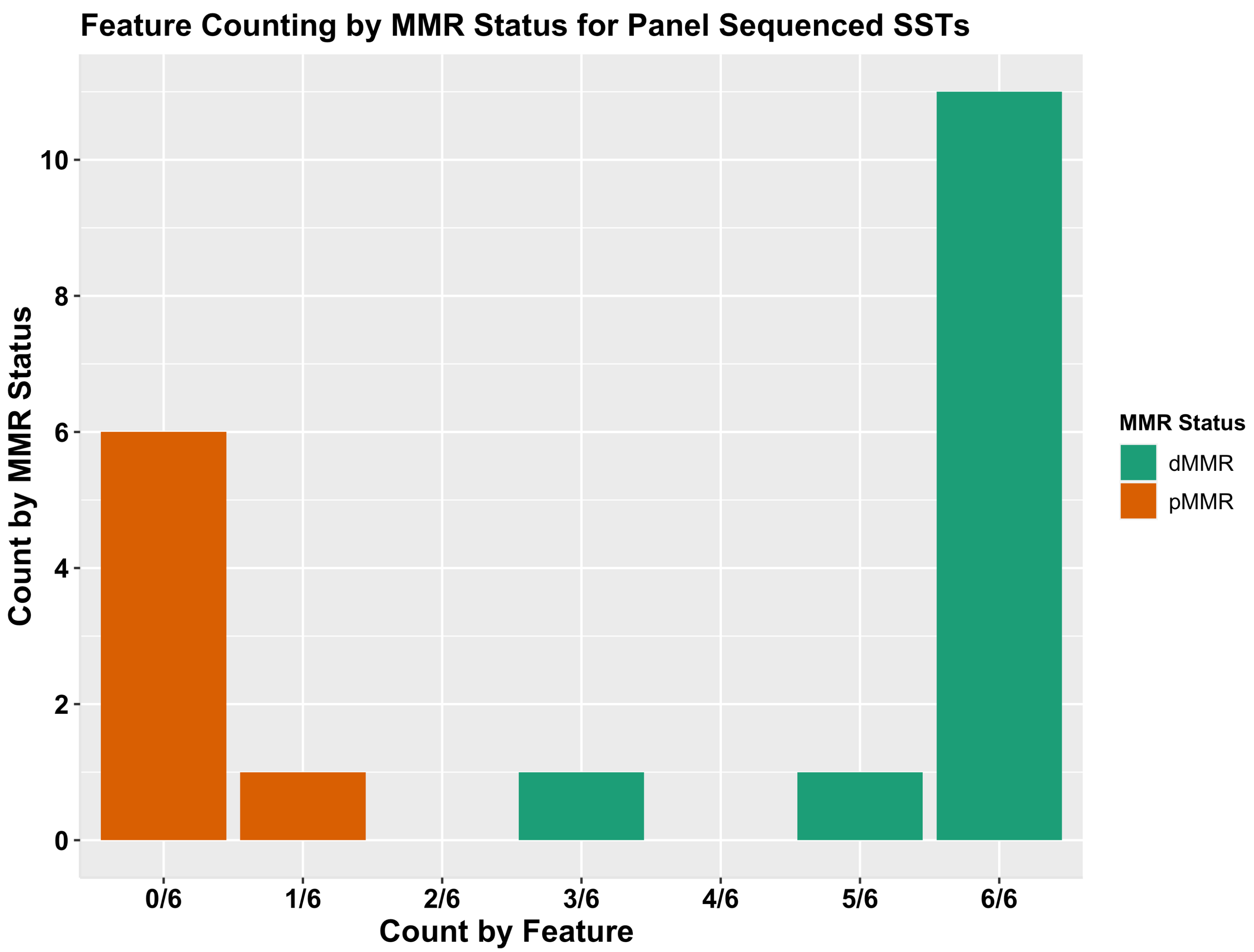
Feature Counting by MMR Status for Panel Sequenced SSTs
Count by MMR Status
0
2
4
6
8
10
0/6
1/6
2/6
3/6
4/6
5/6
6/6
Count by Feature
MMR Status
dMMR
pMMR

**Supplementary Figure 10.** The additive tumor feature combination approach demonstrating the distribution of counts of the top six tumor features by the DNA mismatch repair (MMR) status of the 20 panel sequenced sebaceous skin tumors (SSTs). Bar graphs presenting the distribution of tumors after applying the additive tumor feature combination approach with the recommended thresholds for targeted panel sequenced SSTs using a count of ≥3 out of the top six predictors with >95% accuracy consisting of MSMuTect, MANTIS, MSIseq, MSISensor, INDEL (insertions / deletions) count and TMS (tumor mutational signature) ID2+ID7 (small insertions / deletions) for MMR status calling: MMR-deficient (dMMR) versus (MMR-proficient) pMMR.
