## Supplementary Tables for "Evaluating multiple next-generation sequencing derived tumor features to accurately predict DNA mismatch repair status"

| | CRC WES (n = 300) | |
|---|---|---|
| | **dMMR (n = 91, 30.3%)** | **pMMR (n = 209, 69.7%)** |
| **Gender, n ( %)** | | |
| Male | 34 (37.4%) | 93 (44.5%) |
| Female | 57 (62.6%) | 116 (55.5%) |
| **Age at diagnosis, n ( %)** | | |
| ≤50 years | 50 (54.9%) | 137 (65.6%) |
| >50 years | 41 (45.1%) | 72 (34.4%) |
| Mean ± SD | 50.9 ± 15.0 | 49.0 ± 16.3 |
| Min. - Max. | 18 - 92 | 18 - 89 |
| **Tumor site, n ( %)** | | |
| Proximal | 70 (76.9%) | 72 (34.4%) |
| Distal | 13 (14.3%) | 84 (40.2%) |
| Rectum | 8 (8.8%) | 53 (25.4%) |
| **CRC Histological Type, n ( %)** | | |
| Adenocarcinoma | 70 (76.9%) | 186 (89%) |
| Mucinous | 19 (20.9%) | 5 (2.4%) |
| Signet ring | 1 (1.1%) | 1 (0.5%) |
| Undifferentiated | 1 (1.1%) | 1 (0.5%) |
| Other | 0 (0%) | 4 (1.9%) |
| Unknown | 0 (0%) | 12 (5.7%) |
| **dMMR subtype, n ( %)** | | |
| Lynch syndrome | 49 (53.8%) | - |

| | | |
|---|---|---|
| *MLH1* methylation | 26 (28.6%) | - |
| Double somatic | 16 (17.6%) | - |
| **MMR IHC pattern, n (%)** | | |
| MLH1/PMS2 | 60 (65.9%) | - |
| MSH2/MSH6 | 16 (17.6%) | - |
| MSH6 | 9 (9.9%) | - |
| PMS2 | 6 (6.6%) | - |
| **Study, n ( %)** | | |
| ANGELS | 1 (1.1%) | 61 (29.2%) |
| ACCFR | 64 (70.3%) | 41 (19.6%) |
| OFCCR | 6 (6.6%) | 47 (22.5%) |
| WEHI | 20 (22%) | 60 (28.7%) |

**Supplementary Table 1**. Characteristics of the whole-exome sequenced (WES) colorectal cancers (CRCs). This table shows an overview of the WES CRC cohort by gender, age at diagnosis (including mean and standard deviation, SD), tumor site, histological type, DNA mismatch repair (MMR) deficient (dMMR) subtype, MMR immunohistochemistry pattern and study separated by dMMR and MMR-proficient subgroups.

| | CRC (n = 29) | | EC (n = 22) | | SST (n = 20) | | TOTAL (n = 71) | |
|---|---|---|---|---|---|---|---|---|
| | **dMMR (n = 21, 72.4%)** | **pMMR (n = 8, 27.6%)** | **dMMR (n = 18, 81.8%)** | **pMMR (n = 4, 18.2%)** | **dMMR (n = 13, 65.0%)** | **pMMR (n = 7, 35.0%)** | **dMMR (n = 52, 73.2%)** | **pMMR (n = 19, 26.8%)** |
| **Gender, n ( %)** | | | | | | | | |
| Male | 7 (33.3%) | 4 (50%) | 0 (0%) | 0 (0%) | 11 (84.6%) | 5 (71.4%) | 18 (34.6%) | 9 (47.4%) |
| Female | 14 (66.7%) | 4 (50%) | 18 (100%) | 4 (100%) | 2 (15.4%) | 2 (28.6%) | 34 (65.4%) | 10 (52.6%) |
| **Age at diagnosis, n ( %)** | | | | | | | | |
| Mean ± SD | 49 ± 15.2 | 40 ± 11.7 | 56 ± 10.3 | 73 ± 7.9 | 61 ± 13.2 | 70 ± 8.9 | 54 ± 13.8 | 58 ± 18.5 |
| Min. - Max. | 19 - 79 | 29 - 64 | 40 - 77 | 65 - 83 | 39 - 77 | 59 - 82 | 19 - 79 | 29 - 83 |
| ≤50 years | 11 (52.4%) | 6 (75%) | 6 (33.3%) | 0 (0%) | 3 (23.1%) | 0 (0%) | 20 (38.5%) | 7 (36.8%) |
| >50 years | 10 (47.6%) | 2 (25%) | 12 (66.7%) | 4 (100%) | 10 (76.9%) | 7 (100%) | 32 (61.5%) | 12 (63.2%) |
| **CRC Tumor site, n ( %)** | | | | | | | | |
| Proximal | 17 (81%) | 4 (50%) | - | - | - | - | - | - |
| Distal | 4 (19%) | 2 (25%) | - | - | - | - | - | - |
| Rectum | 0 (0%) | 1 (12.5%) | - | - | - | - | - | - |
| Unknown | 0 (0%) | 1 (12.5%) | - | - | - | - | - | - |
| **CRC Histological Type, n ( %)** | | | | | | | | |
| Adenocarcinoma | 16 (76.2%) | 8 (100%) | - | - | - | - | - | - |
| Mucinous | 5 (23.8%) | 0 (0%) | - | - | - | - | - | - |

| | | | | | | | | |
|---|---|---|---|---|---|---|---|---|
| **EC Histological Type, n ( %)** | | | | | | | | |
| Endometroid | - | - | 16 (100%) | 2 (50%) | - | - | - | - |
| Other | - | - | 0 (0%) | 2 (50%) | - | - | - | - |
| **SST Tumor site, n ( %)** | | | | | | | | |
| Head and neck | - | - | - | - | 7 (53.8%) | 5 (71.4%) | - | - |
| Trunk and limb | - | - | - | - | 6 (46.2%) | 2 (28.6%) | - | - |
| **SST Histological Type, n ( %)** | | | | | | | | |
| Adenoma | - | - | - | - | 11 (84.6%) | 2 (28.6%) | - | - |
| Carcinoma | - | - | - | - | 0 (0%) | 5 (71.4%) | - | - |
| Sebaceoma | - | - | - | - | 1 (7.7%) | 0 (0%) | - | - |
| Unknown | - | - | - | - | 1 (7.7%) | 0 (0%) | - | - |
| **dMMR subtype, n ( %)** | | | | | | | | |
| Lynch syndrome | 14 (66.7%) | - | 6 (33.3%) | - | 6 (46.2%) | - | 26 (50%) | - |
| *MLH1* methylation | 4 (19%) | - | 6 (33.3%) | - | 0 (0%) | - | 10 (19.2%) | - |
| Double somatic | 3 (14.3%) | - | 6 (33.3%) | - | 7 (53.8%) | - | 16 (30.8%) | - |
| **MMR IHC pattern, n (%)** | | | | | | | | |
| MLH1/PMS2 | 10 (47.6%) | - | 9 (50%) | - | 0 (0%) | - | 19 (36.5%) | - |
| MSH2/MSH6 | 6 (28.6%) | - | 7 (38.9%) | - | 10 (76.9%) | - | 23 (44.2%) | - |
| MSH6 | 4 (19%) | - | 2 (11.1%) | - | 3 (23.1%) | - | 9 (17.3%) | - |
| PMS2 | 1 (4.8%) | - | 0 (0%) | - | 0 (0%) | - | 1 (1.9%) | - |
| **Study, n ( %)** | | | | | | | | |

| | | | | | | | | |
|---|---|---|---|---|---|---|---|---|
| ANGELS | 2 (9.5%) | 5 (62.5%) | 9 (50%) | 1 (25%) | 0 (0%) | 0 (0%) | 11 (21.2%) | 6 (31.6%) |
| ACCFR | 19 (90.5%) | 3 (37.5%) | 9 (50%) | 3 (75%) | 0 (0%) | 0 (0%) | 28 (53.8%) | 6 (31.6%) |
| MTS | 0 (0%) | 0 (0%) | 0 (0%) | 0 (0%) | 13 (100%) | 7 (100%) | 13 (25%) | 7 (36.8%) |

**Supplementary Table 2**. Characteristics of the colorectal cancer (CRC), endometrial cancer (EC) and sebaceous skin tumors (SST) with targeted panel sequencing data included in the study. This table shows an overview of the targeted panel sequenced CRCs, ECs and SSTs by gender, age at diagnosis (including mean and standard deviation, SD), tumor site, histological type, DNA mismatch repair (MMR) deficient (dMMR) subtype, MMR immunohistochemistry pattern and study separated by dMMR and MMR-proficient subgroups.

| Categories | Tumor Feature | Mean Accuracy | Error Rate | 95% CI (Accuracy) | Mean Sensitivity | 95% CI (Sensitivity) | Mean Specificity | 95% CI (Specificity) | Mean AUC | 95% CI (AUC) |
|---|---|---|---|---|---|---|---|---|---|---|
| Strong predictor | MSMuTect | 99.3% | 0.7% | 99.1% - 99.5% | 97.6% | 96.9% - 98.3% | 100.0% | 100.0% - 100.0% | 98.8% | 98.5% - 99.1% |
| | MSIseq | 99.1% | 0.9% | 98.9% - 99.4% | 97.7% | 97.0% - 98.3% | 99.8% | 99.6% - 100.0% | 98.7% | 98.4% - 99.1% |
| | MANTIS | 99.0% | 1.0% | 98.8% - 99.2% | 97.1% | 96.4% - 97.7% | 99.9% | 99.8% - 100.0% | 98.5% | 98.1% - 98.8% |
| | INDEL count | 98.9% | 1.1% | 98.7% - 99.2% | 97.7% | 97.0% - 98.3% | 99.5% | 99.2% - 99.8% | 98.6% | 98.2% - 98.9% |
| | MSISensor | 97.7% | 2.3% | 97.3% - 98.0% | 93.4% | 92.4% - 94.5% | 99.5% | 99.3% - 99.7% | 96.5% | 96.0% - 97.0% |
| | TMS ID2+ID7 | 96.8% | 3.2% | 96.4% - 97.2% | 94.2% | 93.2% - 95.2% | 97.9% | 97.5% - 98.4% | 96.0% | 95.5% - 96.6% |
| | TMS ID2 | 93.3% | 6.7% | 92.8% - 93.8% | 90.7% | 89.5% - 91.9% | 94.4% | 93.7% - 95.1% | 92.6% | 92.0% - 93.1% |
| | TMS SBS20 | 88.4% | 11.6% | 87.6% - 89.2% | 68.9% | 66.6% - 71.2% | 97.0% | 96.4% - 97.6% | 82.9% | 81.8% - 84.1% |
| | TMS ID7 | 87.6% | 12.4% | 87.0% - 88.3% | 74.2% | 72.6% - 75.9% | 93.5% | 92.8% - 94.2% | 83.9% | 83.0% - 84.7% |
| | TMS SBS54 | 83.4% | 16.6% | 82.6% - 84.2% | 59.4% | 57.5% - 61.4% | 93.9% | 93.1% - 94.7% | 76.7% | 75.6% - 77.7% |
| | TMB | 83.3% | 16.7% | 82.6% - 83.9% | 57.8% | 55.2% - 60.4% | 94.5% | 93.7% - 95.2% | 76.1% | 75.0% - 77.3% |
| | TMS SBS15 | 82.4% | 17.6% | 81.5% - 83.3% | 58.8% | 56.5% - 61.1% | 92.8% | 91.9% - 93.7% | 75.8% | 74.6% - 77.0% |
| Weak predictor | TMS ID1 | 76.4% | 23.6% | 75.7% - 77.1% | 54.2% | 52.1% - 56.3% | 86.2% | 85.1% - 87.3% | 70.2% | 69.3% - 71.0% |
| | TMS SBS26 | 76.2% | 23.8% | 75.6% - 76.8% | 26.8% | 24.9% - 28.6% | 97.9% | 97.5% - 98.3% | 62.3% | 61.4% - 63.3% |
| | TMS SBS21 | 76.2% | 23.8% | 75.4% - 76.9% | 36.6% | 34.7% - 38.5% | 93.5% | 92.7% - 94.3% | 65.1% | 64.1% - 66.0% |
| | TMS SBS14 | 72.1% | 27.9% | 71.4% - 72.8% | 19.1% | 17.5% - 20.6% | 95.3% | 94.6% - 96.1% | 57.2% | 56.4% - 58.0% |
| | TMS SBS13 | 71.3% | 28.7% | 70.6% - 72.0% | 65.4% | 57.3% - 73.5% | 73.9% | 70.5% - 77.2% | 69.6% | 67.1% - 72.1% |
| | TMS SBS6 | 70.2% | 29.8% | 69.5% - 70.9% | 16.8% | 15.3% - 18.4% | 93.6% | 92.8% - 94.5% | 55.2% | 54.4% - 56.1% |
| | SNV count | 70.2% | 29.8% | 69.7% - 70.6% | 14.4% | 12.5% - 16.4% | 94.6% | 93.9% - 95.3% | 54.5% | 53.7% - 55.3% |

| | | | | | | | | | | |
|---|---|---|---|---|---|---|---|---|---|---|
| | TMS SBS52 | 70.0% | 30.0% | 69.9% - 70.2% | 1.8% | 1.3% - 2.3% | 100.0% | 100.0% - 100.0% | 50.9% | 50.6% - 51.1% |
| | TMS SBS45 | 69.9% | 30.1% | 69.4% - 70.4% | 7.7% | 6.6% - 8.8% | 97.2% | 96.6% - 97.8% | 52.4% | 51.8% - 53.0% |
| Not predictive | TMS ID11 | 69.5% | 30.5% | 69.5% - 69.5% | 0.0% | 0.0% - 0.0% | 100.0% | 100.0% - 100.0% | 50.0% | 50.0% - 50.0% |
| | TMS ID12 | 69.5% | 30.5% | 69.5% - 69.5% | 0.0% | 0.0% - 0.0% | 100.0% | 100.0% - 100.0% | 50.0% | 50.0% - 50.0% |
| | TMS ID13 | 69.5% | 30.5% | 69.5% - 69.5% | 0.0% | 0.0% - 0.0% | 100.0% | 100.0% - 100.0% | 50.0% | 50.0% - 50.0% |
| | TMS ID14 | 69.5% | 30.5% | 69.5% - 69.5% | 0.0% | 0.0% - 0.0% | 100.0% | 100.0% - 100.0% | 50.0% | 50.0% - 50.0% |
| | TMS ID15 | 69.5% | 30.5% | 69.5% - 69.5% | 0.0% | 0.0% - 0.0% | 100.0% | 100.0% - 100.0% | 50.0% | 50.0% - 50.0% |
| | TMS ID16 | 69.5% | 30.5% | 69.5% - 69.5% | 0.0% | 0.0% - 0.0% | 100.0% | 100.0% - 100.0% | 50.0% | 50.0% - 50.0% |
| | TMS ID17 | 69.5% | 30.5% | 69.5% - 69.5% | 0.0% | 0.0% - 0.0% | 100.0% | 100.0% - 100.0% | 50.0% | 50.0% - 50.0% |
| | TMS ID18 | 69.5% | 30.5% | 69.5% - 69.5% | 0.0% | 0.0% - 0.0% | 100.0% | 100.0% - 100.0% | 50.0% | 50.0% - 50.0% |
| | TMS ID3 | 69.5% | 30.5% | 69.5% - 69.5% | 0.0% | 0.0% - 0.0% | 100.0% | 100.0% - 100.0% | 50.0% | 50.0% - 50.0% |
| | TMS ID5 | 69.5% | 30.5% | 69.5% - 69.5% | 0.0% | 0.0% - 0.0% | 100.0% | 100.0% - 100.0% | 50.0% | 50.0% - 50.0% |
| | TMS ID6 | 69.5% | 30.5% | 69.5% - 69.5% | 0.0% | 0.0% - 0.0% | 100.0% | 100.0% - 100.0% | 50.0% | 50.0% - 50.0% |
| | TMS ID8 | 69.5% | 30.5% | 69.5% - 69.5% | 0.0% | 0.0% - 0.0% | 100.0% | 100.0% - 100.0% | 50.0% | 50.0% - 50.0% |
| | TMS ID9 | 69.5% | 30.5% | 69.5% - 69.5% | 0.0% | 0.0% - 0.0% | 100.0% | 100.0% - 100.0% | 50.0% | 50.0% - 50.0% |
| | TMS SBS10a | 69.5% | 30.5% | 69.5% - 69.5% | 0.0% | 0.0% - 0.0% | 100.0% | 100.0% - 100.0% | 50.0% | 50.0% - 50.0% |
| | TMS SBS10b | 69.5% | 30.5% | 69.5% - 69.5% | 0.0% | 0.0% - 0.0% | 100.0% | 100.0% - 100.0% | 50.0% | 50.0% - 50.0% |
| | TMS SBS10c | 69.5% | 30.5% | 69.5% - 69.5% | 0.0% | 0.0% - 0.0% | 100.0% | 100.0% - 100.0% | 50.0% | 50.0% - 50.0% |
| | TMS SBS11 | 69.5% | 30.5% | 69.5% - 69.5% | 0.0% | 0.0% - 0.0% | 100.0% | 100.0% - 100.0% | 50.0% | 50.0% - 50.0% |
| | TMS SBS12 | 69.5% | 30.5% | 69.5% - 69.5% | 0.0% | 0.0% - 0.0% | 100.0% | 100.0% - 100.0% | 50.0% | 50.0% - 50.0% |
| | TMS SBS16 | 69.5% | 30.5% | 69.5% - 69.5% | 0.0% | 0.0% - 0.0% | 100.0% | 100.0% - 100.0% | 50.0% | 50.0% - 50.0% |
| | TMS SBS17a | 69.5% | 30.5% | 69.5% - 69.5% | 0.0% | 0.0% - 0.0% | 100.0% | 100.0% - 100.0% | 50.0% | 50.0% - 50.0% |

| | | | | | | | | | | |
|---|---|---|---|---|---|---|---|---|---|---|
| | TMS SBS17b | 69.5% | 30.5% | 69.5% - 69.5% | 0.0% | 0.0% - 0.0% | 100.0% | 100.0% - 100.0% | 50.0% | 50.0% - 50.0% |
| | TMS SBS18 | 69.5% | 30.5% | 69.5% - 69.5% | 0.0% | 0.0% - 0.0% | 100.0% | 100.0% - 100.0% | 50.0% | 50.0% - 50.0% |
| | TMS SBS19 | 69.5% | 30.5% | 69.5% - 69.5% | 0.0% | 0.0% - 0.0% | 100.0% | 100.0% - 100.0% | 50.0% | 50.0% - 50.0% |
| | TMS SBS2 | 69.5% | 30.5% | 69.5% - 69.5% | 0.0% | 0.0% - 0.0% | 100.0% | 100.0% - 100.0% | 50.0% | 50.0% - 50.0% |
| | TMS SBS22 | 69.5% | 30.5% | 69.5% - 69.5% | 0.0% | 0.0% - 0.0% | 100.0% | 100.0% - 100.0% | 50.0% | 50.0% - 50.0% |
| | TMS SBS23 | 69.5% | 30.5% | 69.5% - 69.5% | 0.0% | 0.0% - 0.0% | 100.0% | 100.0% - 100.0% | 50.0% | 50.0% - 50.0% |
| | TMS SBS24 | 69.5% | 30.5% | 69.5% - 69.5% | 0.0% | 0.0% - 0.0% | 100.0% | 100.0% - 100.0% | 50.0% | 50.0% - 50.0% |
| | TMS SBS25 | 69.5% | 30.5% | 69.5% - 69.5% | 0.0% | 0.0% - 0.0% | 100.0% | 100.0% - 100.0% | 50.0% | 50.0% - 50.0% |
| | TMS SBS27 | 69.5% | 30.5% | 69.5% - 69.5% | 0.0% | 0.0% - 0.0% | 100.0% | 100.0% - 100.0% | 50.0% | 50.0% - 50.0% |
| | TMS SBS28 | 69.5% | 30.5% | 69.5% - 69.5% | 0.0% | 0.0% - 0.0% | 100.0% | 100.0% - 100.0% | 50.0% | 50.0% - 50.0% |
| | TMS SBS29 | 69.5% | 30.5% | 69.5% - 69.5% | 0.0% | 0.0% - 0.0% | 100.0% | 100.0% - 100.0% | 50.0% | 50.0% - 50.0% |
| | TMS SBS3 | 69.5% | 30.5% | 69.5% - 69.5% | 0.0% | 0.0% - 0.0% | 100.0% | 100.0% - 100.0% | 50.0% | 50.0% - 50.0% |
| | TMS SBS30 | 69.5% | 30.5% | 69.5% - 69.5% | 0.0% | 0.0% - 0.0% | 100.0% | 100.0% - 100.0% | 50.0% | 50.0% - 50.0% |
| | TMS SBS31 | 69.5% | 30.5% | 69.5% - 69.5% | 0.0% | 0.0% - 0.0% | 100.0% | 100.0% - 100.0% | 50.0% | 50.0% - 50.0% |
| | TMS SBS32 | 69.5% | 30.5% | 69.5% - 69.5% | 0.0% | 0.0% - 0.0% | 100.0% | 100.0% - 100.0% | 50.0% | 50.0% - 50.0% |
| | TMS SBS33 | 69.5% | 30.5% | 69.5% - 69.5% | 0.0% | 0.0% - 0.0% | 100.0% | 100.0% - 100.0% | 50.0% | 50.0% - 50.0% |
| | TMS SBS34 | 69.5% | 30.5% | 69.5% - 69.5% | 0.0% | 0.0% - 0.0% | 100.0% | 100.0% - 100.0% | 50.0% | 50.0% - 50.0% |
| | TMS SBS35 | 69.5% | 30.5% | 69.5% - 69.5% | 0.0% | 0.0% - 0.0% | 100.0% | 100.0% - 100.0% | 50.0% | 50.0% - 50.0% |

| | | | | | | | | | | |
|---|---|---|---|---|---|---|---|---|---|---|
| | TMS SBS36 | 69.5% | 30.5% | 69.5% - 69.5% | 0.0% | 0.0% - 0.0% | 100.0% | 100.0% - 100.0% | 50.0% | 50.0% - 50.0% |
| | TMS SBS38 | 69.5% | 30.5% | 69.5% - 69.5% | 0.0% | 0.0% - 0.0% | 100.0% | 100.0% - 100.0% | 50.0% | 50.0% - 50.0% |
| | TMS SBS39 | 69.5% | 30.5% | 69.5% - 69.5% | 0.0% | 0.0% - 0.0% | 100.0% | 100.0% - 100.0% | 50.0% | 50.0% - 50.0% |
| | TMS SBS4 | 69.5% | 30.5% | 69.5% - 69.5% | 0.0% | 0.0% - 0.0% | 100.0% | 100.0% - 100.0% | 50.0% | 50.0% - 50.0% |
| | TMS SBS40 | 69.5% | 30.5% | 69.5% - 69.5% | 0.0% | 0.0% - 0.0% | 100.0% | 100.0% - 100.0% | 50.0% | 50.0% - 50.0% |
| | TMS SBS41 | 69.5% | 30.5% | 69.5% - 69.5% | 0.0% | 0.0% - 0.0% | 100.0% | 100.0% - 100.0% | 50.0% | 50.0% - 50.0% |
| | TMS SBS42 | 69.5% | 30.5% | 69.5% - 69.5% | 0.0% | 0.0% - 0.0% | 100.0% | 100.0% - 100.0% | 50.0% | 50.0% - 50.0% |
| | TMS SBS44 | 69.5% | 30.5% | 69.5% - 69.5% | 0.0% | 0.0% - 0.0% | 100.0% | 100.0% - 100.0% | 50.0% | 50.0% - 50.0% |
| | TMS SBS47 | 69.5% | 30.5% | 69.5% - 69.5% | 0.0% | 0.0% - 0.0% | 100.0% | 100.0% - 100.0% | 50.0% | 50.0% - 50.0% |
| | TMS SBS48 | 69.5% | 30.5% | 69.5% - 69.5% | 0.0% | 0.0% - 0.0% | 100.0% | 100.0% - 100.0% | 50.0% | 50.0% - 50.0% |
| | TMS SBS5 | 69.5% | 30.5% | 69.5% - 69.5% | 0.0% | 0.0% - 0.0% | 100.0% | 100.0% - 100.0% | 50.0% | 50.0% - 50.0% |
| | TMS SBS50 | 69.5% | 30.5% | 69.5% - 69.5% | 0.0% | 0.0% - 0.0% | 100.0% | 100.0% - 100.0% | 50.0% | 50.0% - 50.0% |
| | TMS SBS51 | 69.5% | 30.5% | 69.5% - 69.5% | 0.0% | 0.0% - 0.0% | 100.0% | 100.0% - 100.0% | 50.0% | 50.0% - 50.0% |
| | TMS SBS53 | 69.5% | 30.5% | 69.5% - 69.5% | 0.0% | 0.0% - 0.0% | 100.0% | 100.0% - 100.0% | 50.0% | 50.0% - 50.0% |
| | TMS SBS55 | 69.5% | 30.5% | 69.5% - 69.5% | 0.0% | 0.0% - 0.0% | 100.0% | 100.0% - 100.0% | 50.0% | 50.0% - 50.0% |
| | TMS SBS56 | 69.5% | 30.5% | 69.5% - 69.5% | 0.0% | 0.0% - 0.0% | 100.0% | 100.0% - 100.0% | 50.0% | 50.0% - 50.0% |
| | TMS SBS58 | 69.5% | 30.5% | 69.5% - 69.5% | 0.0% | 0.0% - 0.0% | 100.0% | 100.0% - 100.0% | 50.0% | 50.0% - 50.0% |
| | TMS SBS59 | 69.5% | 30.5% | 69.5% - 69.5% | 0.0% | 0.0% - 0.0% | 100.0% | 100.0% - 100.0% | 50.0% | 50.0% - 50.0% |

| | | | | | | | | | | |
|---|---|---|---|---|---|---|---|---|---|---|
| | TMS SBS60 | 69.5% | 30.5% | 69.5% - 69.5% | 0.0% | 0.0% - 0.0% | 100.0% | 100.0% - 100.0% | 50.0% | 50.0% - 50.0% |
| | TMS SBS7a | 69.5% | 30.5% | 69.5% - 69.5% | 0.0% | 0.0% - 0.0% | 100.0% | 100.0% - 100.0% | 50.0% | 50.0% - 50.0% |
| | TMS SBS7b | 69.5% | 30.5% | 69.5% - 69.5% | 0.0% | 0.0% - 0.0% | 100.0% | 100.0% - 100.0% | 50.0% | 50.0% - 50.0% |
| | TMS SBS7d | 69.5% | 30.5% | 69.5% - 69.5% | 0.0% | 0.0% - 0.0% | 100.0% | 100.0% - 100.0% | 50.0% | 50.0% - 50.0% |
| | TMS SBS8 | 69.5% | 30.5% | 69.5% - 69.5% | 0.0% | 0.0% - 0.0% | 100.0% | 100.0% - 100.0% | 50.0% | 50.0% - 50.0% |
| | TMS SBS84 | 69.5% | 30.5% | 69.5% - 69.5% | 0.0% | 0.0% - 0.0% | 100.0% | 100.0% - 100.0% | 50.0% | 50.0% - 50.0% |
| | TMS SBS85 | 69.5% | 30.5% | 69.5% - 69.5% | 0.0% | 0.0% - 0.0% | 100.0% | 100.0% - 100.0% | 50.0% | 50.0% - 50.0% |
| | TMS SBS86 | 69.5% | 30.5% | 69.5% - 69.5% | 0.0% | 0.0% - 0.0% | 100.0% | 100.0% - 100.0% | 50.0% | 50.0% - 50.0% |
| | TMS SBS87 | 69.5% | 30.5% | 69.5% - 69.5% | 0.0% | 0.0% - 0.0% | 100.0% | 100.0% - 100.0% | 50.0% | 50.0% - 50.0% |
| | TMS SBS88 | 69.5% | 30.5% | 69.5% - 69.5% | 0.0% | 0.0% - 0.0% | 100.0% | 100.0% - 100.0% | 50.0% | 50.0% - 50.0% |
| | TMS SBS89 | 69.5% | 30.5% | 69.5% - 69.5% | 0.0% | 0.0% - 0.0% | 100.0% | 100.0% - 100.0% | 50.0% | 50.0% - 50.0% |
| | TMS SBS9 | 69.5% | 30.5% | 69.5% - 69.5% | 0.0% | 0.0% - 0.0% | 100.0% | 100.0% - 100.0% | 50.0% | 50.0% - 50.0% |
| | TMS SBS90 | 69.5% | 30.5% | 69.5% - 69.5% | 0.0% | 0.0% - 0.0% | 100.0% | 100.0% - 100.0% | 50.0% | 50.0% - 50.0% |
| | TMS SBS91 | 69.5% | 30.5% | 69.5% - 69.5% | 0.0% | 0.0% - 0.0% | 100.0% | 100.0% - 100.0% | 50.0% | 50.0% - 50.0% |
| | TMS SBS92 | 69.5% | 30.5% | 69.5% - 69.5% | 0.0% | 0.0% - 0.0% | 100.0% | 100.0% - 100.0% | 50.0% | 50.0% - 50.0% |
| | TMS SBS93 | 69.5% | 30.5% | 69.5% - 69.5% | 0.0% | 0.0% - 0.0% | 100.0% | 100.0% - 100.0% | 50.0% | 50.0% - 50.0% |
| | TMS SBS94 | 69.5% | 30.5% | 69.5% - 69.5% | 0.0% | 0.0% - 0.0% | 100.0% | 100.0% - 100.0% | 50.0% | 50.0% - 50.0% |
| | TMS SBS10d | 69.5% | 30.5% | 69.4% - 69.5% | 0.0% | 0.0% - 0.0% | 100.0% | 99.9% - 100.0% | 50.0% | 50.0% - 50.0% |

| | | | | | | | | | | |
|---|---|---|---|---|---|---|---|---|---|---|
| | TMS SBS43 | 69.5% | 30.5% | 69.4% - 69.5% | 0.1% | 0.0% - 0.3% | 99.9% | 99.8% - 100.0% | 50.0% | 49.9% - 50.1% |
| | TMS SBS1 | 69.4% | 30.6% | 69.4% - 69.5% | 0.0% | 0.0% - 0.0% | 99.9% | 99.8% - 100.0% | 50.0% | 49.9% - 50.0% |
| | TMS SBS37 | 69.4% | 30.6% | 69.3% - 69.5% | 0.0% | 0.0% - 0.0% | 99.9% | 99.8% - 100.1% | 50.0% | 49.9% - 50.0% |
| | TMS SBS7c | 69.4% | 30.6% | 69.3% - 69.5% | 0.0% | 0.0% - 0.0% | 99.9% | 99.7% - 100.0% | 49.9% | 49.9% - 50.0% |
| | TMS SBS46 | 69.4% | 30.6% | 68.9% - 69.9% | 6.6% | 5.6% - 7.5% | 96.9% | 96.3% - 97.5% | 51.7% | 51.2% - 52.3% |
| | TMS SBS49 | 69.2% | 30.8% | 69.0% - 69.4% | 0.0% | 0.0% - 0.0% | 99.6% | 99.3% - 99.9% | 49.8% | 49.7% - 49.9% |
| | TMS SBS57 | 69.1% | 30.9% | 68.6% - 69.6% | 6.9% | 5.9% - 8.0% | 96.4% | 95.8% - 97.0% | 51.7% | 51.1% - 52.2% |
| | TMS ID10 | 68.4% | 31.6% | 67.7% - 69.0% | 13.4% | 6.9% - 20.0% | 92.5% | 88.8% - 96.2% | 53.0% | 51.5% - 54.4% |
| | TMS ID4 | 66.9% | 33.1% | 66.0% - 67.8% | 27.7% | 21.0% - 34.4% | 84.1% | 80.3% - 88.0% | 55.9% | 54.3% - 57.5% |

**Supplementary Table 3.** Performance of the 104 tumor features from whole-exome sequenced (WES) colorectal cancers (CRCs) using 10-fold cross validated analysis with 100 repeats. Table presents the mean accuracy after 10-fold cross-validation with 100 repeats, error rate, mean sensitivity, mean specificity, and mean area under the curves (AUCs) with corresponding 95% confidence intervals (CIs) for the 104 tumor features from the WES CRC analysis. INDEL, insertions / deletions; TMB, tumor mutation burden; TMS, tumor mutational signature; ID, small insertions, and deletions; SBS, single base substitutions; SNV, single nucleotide variant count.

| | MMR Status | | | | dMMR Subtype | | | |
|---|---|---|---|---|---|---|---|---|
| | pMMR (n = 209, [69.7%]) | dMMR (n = 91, [30.3%]) | t-test (p-value)* | Effect Size (Cohen's d) | dMMR-Lynch (n = 49 [16.3%]) | dMMR-MLH1me (n = 26 [8.7%]) | dMMR-DS (n = 16, [5.3%]) | Total (n = 300, [100%]) |
| MSMuTect | | | **6.45913E-37** | 4.0 | | | | |
| Mean | 353.6 | 3571.4 | | | 3703.9 | 3171.6 | 3814.9 | 1329.6 |
| SD | 143.4 | 1445.1 | | | 1364.4 | 1469.9 | 1604.2 | 1684.7 |
| Range | 33-754 | 70-7093 | | | 1065-7093 | 70-5442 | 1254-6436 | 33-7093 |
| MSIseq | | | **6.87604E-32** | 3.5 | | | | |
| Mean | 0.3 | 28.8 | | | 29.3 | 26.2 | 31.5 | 8.9 |
| SD | 0.3 | 15.0 | | | 16.1 | 13.3 | 14.1 | 15.5 |
| Range | 0.0-2.8 | 0.1-73.7 | | | 5.0-73.7 | 0.1-55.9 | 8.5-60.2 | 0.0-73.7 |
| MANTIS | | | **1.1014E-36** | 4.0 | | | | |
| Mean | 0.2 | 0.6 | | | 0.5 | 0.7 | 0.6 | 0.3 |
| SD | 0.0 | 0.2 | | | 0.2 | 0.2 | 0.1 | 0.2 |
| Range | 0.160-0.238 | 0.179-1.020 | | | 0.245-0.773 | 0.179-1.020 | 0.337-0.877 | 0.160-1.020 |
| INDEL count | | | **7.3226E-32** | 3.5 | | | | |
| Mean | 33.6 | 2137.8 | | | 2161.3 | 1964.2 | 2348.3 | 671.9 |
| SD | 33.5 | 1105.0 | | | 1197.8 | 975.0 | 1024.6 | 1143.3 |
| Range | 7-269 | 16-5531 | | | 352-5531 | 16-4098 | 631-4457 | 7-5531 |
| MSISensor | | | **1.39619E-40** | 4.5 | | | | |
| Mean | 0.9 | 28.7 | | | 30.1 | 25.1 | 30.2 | 9.3 |
| SD | 1.2 | 11.2 | | | 10.5 | 12.6 | 10.0 | 14.2 |

| | | | | | | | | |
|---|---|---|---|---|---|---|---|---|
| Range | 0.0-12.6 | 0.0-46.7 | | | 1.7-46.0 | 0.0-46.7 | 12.1-44.3 | 0.0-46.7 |
| TMS ID2+ID7 | | | **7.77482E-98** | 3.5 | | | | |
| Mean | 0.3 | 0.8 | | | 0.8 | 0.8 | 0.8 | 0.5 |
| SD | 0.1 | 0.1 | | | 0.1 | 0.1 | 0.1 | 0.2 |
| Range | 0.005-0.709 | 0.286-0.925 | | | 0.571-0.908 | 0.286-0.909 | 0.589-0.925 | 0.005-0.925 |
| TMS SBS20 | | | **6.4587E-15** | 1.7 | | | | |
| Mean | 0.0 | 0.1 | | | 0.0 | 0.1 | 0.1 | 0.0 |
| SD | 0.0 | 0.1 | | | 0.0 | 0.1 | 0.1 | 0.0 |
| Range | 0.000-0.082 | 0.000-0.450 | | | 0.000-0.150 | 0.000-0.157 | 0.000-0.450 | 0.000-0.450 |
| TMS SBS54 | | | **4.8626E-15** | 1.5 | | | | |
| Mean | 0.0 | 0.1 | | | 0.1 | 0.1 | 0.1 | 0.0 |
| SD | 0.0 | 0.0 | | | 0.0 | 0.1 | 0.0 | 0.0 |
| Range | 0.000-0.141 | 0.002-0.384 | | | 0.002-0.182 | 0.012-0.384 | 0.013-0.116 | 0.000-0.384 |
| TMB | | | **1.93008E-12** | 1.2 | | | | |
| Mean | 15.9 | 88.1 | | | 82.2 | 84.3 | 112.2 | 37.8 |
| SD | 48.9 | 81.5 | | | 49.9 | 115.2 | 94.7 | 69.0 |
| Range | 0.4-410.1 | 4.2-629.4 | | | 19.2-273.5 | 4.2-629.4 | 19.7-401.6 | 0.4-629.4 |
| TMS SBS15 | | | **4.93144E-16** | 1.5 | | | | |
| Mean | 0.1 | 0.2 | | | 0.2 | 0.3 | 0.2 | 0.1 |
| SD | 0.1 | 0.2 | | | 0.1 | 0.2 | 0.2 | 0.1 |
| Range | 0.000-0.324 | 0.000-0.783 | | | 0.000-0.484 | 0.000-0.709 | 0.000-0.783 | 0.000-0.783 |

**Supplementary Table 4.** The mean, standard deviation, and range of each of the top 10 tumor features stratified by DNA mismatch repair (MMR) proficient (pMMR) and MMR-deficient (dMMR) status including the three dMMR subtypes (dMMR-Lynch, dMMR-MLH1me and dMMR-DS) from the whole exome sequencing (WES) analysis of colorectal cancers (CRCs). Statistically significant p-values are highlighted in **bold**. dMMR-Lynch, Lynch syndrome; dMMR-MLH1me, *MLH1* methylated; dMMR-DS, double somatic MMR mutation; SD, standard deviation; TMS, tumor mutational signature; * heteroscedastic, two-tailed t-test.

| | CRC | CRC | EC | SST |
|---|---|---|---|---|
| | WES | PANEL | PANEL | PANEL |
| Feature | pMMR < n ≥ dMMR | pMMR < n ≥ dMMR | pMMR < n ≥ dMMR | pMMR < n ≥ dMMR |
| MSMuTect | 1065 | 48 | 31 | 25 |
| MSIseq | 2.987 | 2.494 | 0.499 | 0.499 |
| MANTIS | 0.2447 | 0.252 | 0.274 | 0.217 |
| INDEL count | 228 | 5 | 1 | 3 |
| MSISensor | 5.41 | 6.9 | 21.21 | 2.05 |
| TMS ID2+ID7 | 0.605 | 0.504 | 0.182 | 0.547 |
| TMS SBS20 | 0.011 | 0.001 | 0.011 | 0.016 |
| TMS SBS54 | 0.038 | 0.011 | 0.025 | 0.001 |
| TMB | 15.5 | 20.4 | 31.4 | 20.4 |
| TMS SBS15 | 0.224 | 0.123 | 0.099 | 0.034 |

**Supplementary Table 5**. List of recommended thresholds for the top 10 performing features for whole exome sequenced (WES) colorectal cancers (CRCs) and panel sequenced CRC, endometrial cancers (EC) and sebaceous skin tumors (SST) test sets. dMMR, DNA mismatch repair-deficient; pMMR, DNA mismatch repair proficient.

| (A) Colorectal cancer | | | | | | | | |
|---|---|---|---|---|---|---|---|---|
| | MMR Status | | | | dMMR Subtype | | | |
| | pMMR (n = 8, [27.6%]) | dMMR (n = 21, [72.4%]) | t-test (p-value)* | Effect Size (Cohen's d) | dMMR-Lynch (n = 14 [48.3%]) | dMMR-MLH1me (n = 4 [13.8%]) | dMMR-DS (n = 3, [10.3%]) | Total (n = 29, [100%]) |
| MSMuTect | | | **4.4089E-11** | 3.2 | | | | |
| Mean | 18.6 | 190.8 | | | 191.8 | 181.0 | 199.0 | 143.3 |
| SD | 4.6 | 62.5 | | | 75.6 | 19.2 | 34.0 | 94.5 |
| Range | 12-25 | 48-310 | | | 48-310 | 158-204 | 165-233 | 12-310 |
| MSIseq | | | **0.00129168** | 0.9 | | | | |
| Mean | 0.1 | 14.6 | | | 17.3 | 11.1 | 6.5 | 10.6 |
| SD | 0.2 | 17.7 | | | 20.4 | 12.4 | 3.1 | 16.3 |
| Range | 0.0-0.5 | 0.0-76.3 | | | 0.0-76.3 | 2.5-29.4 | 3.0-9.0 | 0.0-76.3 |
| MANTIS | | | **1.6518E-09** | 2.5 | | | | |
| Mean | 0.2 | 0.5 | | | 0.5 | 0.5 | 0.5 | 0.4 |
| SD | 0.0 | 0.1 | | | 0.2 | 0.1 | 0.1 | 0.2 |
| Range | 0.171-0.246 | 0.252-0.939 | | | 0.252-0.939 | 0.442-0.588 | 0.450-0.613 | 0.171-0.939 |
| INDEL count | | | **0.00140578** | 0.9 | | | | |
| Mean | 0.9 | 32.0 | | | 37.7 | 25.5 | 14.3 | 23.4 |
| SD | 1.1 | 38.6 | | | 44.6 | 27.3 | 7.0 | 35.5 |
| Range | 0-3 | 0-167 | | | 0-167 | 5-65 | 7-21 | 0-167 |
| MSISensor | | | **1.4999E-10** | 3.0 | | | | |

| | | | | | | | | |
|---|---|---|---|---|---|---|---|---|
| Mean | 2.3 | 27.5 | | | 27.9 | 24.0 | 30.0 | 20.5 |
| SD | 1.1 | 9.9 | | | 12.0 | 1.1 | 3.2 | 14.2 |
| Range | 1.2-4.1 | 6.9-46.8 | | | 6.9-46.8 | 22.8-25.3 | 27.5-33.6 | 1.2-46.8 |
| TMS ID2+ID7 | | | **0.00113873** | 2.1 | | | | |
| Mean | 0.2 | 0.7 | | | 0.7 | 0.8 | 0.7 | 0.6 |
| SD | 0.3 | 0.2 | | | 0.3 | 0.2 | 0.0 | 0.4 |
| Range | 0.000-0.966 | 0.112-1.000 | | | 0.112-1.000 | 0.504-0.900 | 0.700-0.786 | 0.000-1.000 |
| TMS SBS20 | | | **0.0085012** | 0.8 | | | | |
| Mean | 0.0 | 0.1 | | | 0.0 | 0.2 | 0.0 | 0.1 |
| SD | 0.0 | 0.1 | | | 0.1 | 0.1 | 0.0 | 0.1 |
| Range | 0.000-0.071 | 0.000-0.338 | | | 0.000-0.151 | 0.026-0.338 | 0.000-0.086 | 0.000-0.338 |
| TMS SBS54 | | | **0.00027136** | 1.1 | | | | |
| Mean | 0.0 | 0.1 | | | 0.0 | 0.1 | 0.0 | 0.0 |
| SD | 0.0 | 0.1 | | | 0.0 | 0.1 | 0.0 | 0.0 |
| Range | 0.000-0.000 | 0.000-0.180 | | | 0.000-0.160 | 0.000-0.180 | 0.000-0.059 | 0.000-0.180 |
| TMB | | | **3.5364E-08** | 2.1 | | | | |
| Mean | 7.5 | 58.5 | | | 59.7 | 55.4 | 57.4 | 44.5 |
| SD | 3.7 | 27.6 | | | 32.0 | 23.0 | 10.2 | 32.9 |
| Range | 4.0-14.0 | 5.5-134.2 | | | 5.5-134.2 | 34.9-82.8 | 46.9-67.3 | 4.0-134.2 |
| TMS SBS15 | | | 0.35775826 | 0.4 | | | | |
| Mean | 0.2 | 0.2 | | | 0.2 | 0.1 | 0.2 | 0.2 |
| SD | 0.1 | 0.2 | | | 0.2 | 0.1 | 0.2 | 0.2 |

| | | | | | | | | |
|---|---|---|---|---|---|---|---|---|
| Range | 0.000-0.341 | 0.000-0.252 | | | 0.000-0.631 | 0.000-0.259 | 0.083-0.489 | 0.000-0.631 |
| (B) Endometrial cancer | | | | | | | | |
| | MMR Status | | | | dMMR Subtype | | | |
| | pMMR (n = 4, [18.2%]) | dMMR (n = 18, [81.8%]) | t-test (p-value)* | Effect Size (Cohen's d) | dMMR-Lynch (n = 6 [27.3%]) | dMMR-MLH1me (n = 6 [27.3%]) | dMMR-DS (n = 6, [27.3%]) | Total (n = 22, [100%]) |
| MSMuTect | | | **8.0279E-07** | 1.9 | | | | |
| Mean | 14.3 | 125.3 | | | 145.8 | 96.3 | 133.8 | 105.1 |
| SD | 6.0 | 63.2 | | | 25.3 | 68.9 | 81.3 | 71.9 |
| Range | 10-23 | 20-217 | | | 99-168 | 20-174 | 31-217 | 10-217 |
| MSIseq | | | **0.00123083** | 1.0 | | | | |
| Mean | 0.0 | 8.4 | | | 6.7 | 4.5 | 13.9 | 6.8 |
| SD | 0.0 | 9.2 | | | 6.2 | 7.4 | 11.6 | 8.9 |
| Range | 0.0-0.0 | 0.0-26.9 | | | 1.0-19.0 | 0.0-19.0 | 1.5-26.9 | 0.0-26.9 |
| MANTIS | | | **3.8576E-06** | 1.7 | | | | |
| Mean | 0.2 | 0.5 | | | 0.5 | 0.4 | 0.5 | 0.4 |
| SD | 0.0 | 0.2 | | | 0.1 | 0.2 | 0.2 | 0.2 |
| Range | 0.216-0.266 | 0.217-0.719 | | | 0.400-0.670 | 0.217-0.719 | 0.274-0.636 | 0.216-0.719 |
| INDEL count | | | **0.00061607** | 1.1 | | | | |
| Mean | 0.0 | 19.8 | | | 16.2 | 12.3 | 30.8 | 16.2 |

| | | | | | | | | |
|---|---|---|---|---|---|---|---|---|
| SD | 0.0 | 20.0 | | | 12.1 | 18.4 | 25.5 | 19.6 |
| Range | 0-0 | 0-59 | | | 3-39 | 0-48 | 3-59 | 0-59 |
| MSISensor | | | 0.08571803 | 1.1 | | | | |
| Mean | 14.6 | 24.5 | | | 24.5 | 23.2 | 25.8 | 22.7 |
| SD | 8.2 | 9.4 | | | 7.3 | 14.6 | 5.6 | 9.8 |
| Range | 2.9-21.1 | 2.9-36.1 | | | 13.8-32.8 | 2.9-36.1 | 17.1-30.6 | 2.9-36.1 |
| TMS ID2+ID7 | | | **5.8286E-06** | 1.7 | | | | |
| Mean | 0.1 | 0.6 | | | 0.7 | 0.4 | 0.8 | 0.5 |
| SD | 0.0 | 0.3 | | | 0.3 | 0.4 | 0.2 | 0.4 |
| Range | 0.112-0.112 | 0.000-0.965 | | | 0.182-0.923 | 0.000-0.869 | 0.486-0.965 | 0.000-0.965 |
| TMS SBS20 | | | **0.01729237** | 0.7 | | | | |
| Mean | 0.0 | 0.0 | | | 0.0 | 0.0 | 0.1 | 0.0 |
| SD | 0.0 | 0.1 | | | 0.0 | 0.1 | 0.1 | 0.1 |
| Range | 0.000-0.000 | 0.000-0.267 | | | 0.000-0.067 | 0.000-0.123 | 0.000-0.267 | 0.000-0.267 |
| TMS SBS54 | | | **0.00850301** | 0.8 | | | | |
| Mean | 0.0 | 0.0 | | | 0.0 | 0.0 | 0.0 | 0.0 |
| SD | 0.0 | 0.0 | | | 0.0 | 0.0 | 0.0 | 0.0 |
| Range | 0.000-0.013 | 0.000-0.084 | | | 0.000-0.068 | 0.000-0.034 | 0.000-0.084 | 0.000-0.084 |
| TMB | | | 0.793896 | 0.1 | | | | |
| Mean | 67.3 | 77.8 | | | 55.9 | 50.2 | 127.1 | 75.9 |
| SD | 63.7 | 88.8 | | | 18.3 | 73.2 | 129.3 | 83.6 |
| Range | 15.0-156.1 | 3.0-355.1 | | | 36.9-84.8 | 3.0-197.0 | 37.4-355.1 | 3.0-355.1 |

| | | | | | | | | |
|---|---|---|---|---|---|---|---|---|
| TMS SBS15 | | | **0.03497383** | 0.6 | | | | |
| Mean | 0.0 | 0.1 | | | 0.1 | 0.1 | 0.1 | 0.1 |
| SD | 0.0 | 0.1 | | | 0.1 | 0.2 | 0.1 | 0.1 |
| Range | 0.000-0.017 | 0.000-0.445 | | | 0.000-0.185 | 0.000-0.445 | 0.000-0.217 | 0.000-0.445 |
| (C) Sebaceous skin tumor | | | | | | | | |
| | MMR Status | | | | dMMR Subtype | | | |
| | pMMR (n = 7, [35.0%]) | dMMR (n = 13, [65.0%]) | t-test (p-value)* | Effect Size (Cohen's d) | dMMR-Lynch (n = 6 [30.0%]) | dMMR-MLH1me (n = 0 [0.0%]) | dMMR-DS (n = 7, [35.0%]) | Total (n = 20, [100%]) |
| MSMuTect | | | **0.00020992** | 1.8 | | | | |
| Mean | 12.9 | 89.5 | | | 72.2 | - | 104.3 | 62.7 |
| SD | 4.7 | 53.0 | | | 67.0 | - | 36.3 | 56.4 |
| Range | 5-21 | 25-205 | | | 25-205 | - | 63-174 | 5-205 |
| MSIseq | | | **0.04419683** | 0.8 | | | | |
| Mean | 0.0 | 6.8 | | | 7.7 | - | 5.9 | 4.4 |
| SD | 0.0 | 10.8 | | | 16.0 | - | 4.3 | 9.2 |
| Range | 0.0-0.0 | 0.0-40.4 | | | 0.0-40.4 | - | 0.5-12.0 | 0.0-40.4 |
| MANTIS | | | **0.00195894** | 1.3 | | | | |
| Mean | 0.2 | 0.4 | | | 0.3 | - | 0.4 | 0.3 |
| SD | 0.0 | 0.1 | | | 0.1 | - | 0.1 | 0.1 |
| Range | 0.173-0.273 | 0.217-0.696 | | | 0.217-0.621 | - | 0.270-0.696 | 0.173-0.696 |

| | | | | | | | | |
|---|---|---|---|---|---|---|---|---|
| INDEL count | | | **0.04093278** | 0.8 | | | | |
| Mean | 0.3 | 15.0 | | | 17.0 | - | 13.3 | 9.9 |
| SD | 0.5 | 23.2 | | | 34.3 | - | 9.1 | 19.8 |
| Range | 0-1 | 0-87 | | | 0-87 | - | 1-25 | 0-87 |
| MSISensor | | | **0.00041493** | 1.6 | | | | |
| Mean | 1.0 | 13.8 | | | 10.5 | - | 16.6 | 9.3 |
| SD | 0.6 | 9.6 | | | 11.8 | - | 6.8 | 9.9 |
| Range | 0.0-1.8 | 2.1-34.3 | | | 2.1-34.3 | - | 8.2-27.3 | 0.0-34.3 |
| TMS ID2+ID7 | | | **1.7633E-07** | 3.3 | | | | |
| Mean | 0.1 | 0.8 | | | 0.7 | - | 0.9 | 0.5 |
| SD | 0.1 | 0.3 | | | 0.3 | - | 0.2 | 0.4 |
| Range | 0.000-0.112 | 0.112-1.000 | | | 0.112-1.000 | - | 0.587-1.000 | 0.000-1.000 |
| TMS SBS20 | | | 0.26524707 | 0.4 | | | | |
| Mean | 0.0 | 0.0 | | | 0.0 | - | 0.0 | 0.0 |
| SD | 0.0 | 0.0 | | | 0.0 | - | 0.0 | 0.0 |
| Range | 0.000-0.029 | 0.000-0.110 | | | 0.000-0.110 | - | 0.000-0.032 | 0.000-0.110 |
| TMS SBS54 | | | 0.66664554 | 0.3 | | | | |
| Mean | 0.0 | 0.0 | | | 0.0 | - | 0.0 | 0.0 |
| SD | 0.1 | 0.0 | | | 0.0 | - | 0.0 | 0.1 |
| Range | 0.000-0.217 | 0.000-0.093 | | | 0.000-0.093 | - | 0.000-0.055 | 0.000-0.217 |
| TMB | | | 0.42374512 | 0.5 | | | | |
| Mean | 61.4 | 41.0 | | | 43.5 | - | 38.8 | 48.1 |

| | | | | | | | | |
|---|---|---|---|---|---|---|---|---|
| SD | 60.6 | 27.7 | | | 40.6 | - | 12.4 | 41.8 |
| Range | 2.5-146.6 | 7.0-97.8 | | | 7.0-97.8 | - | 20.4-52.9 | 2.5-146.6 |
| TMS SBS15 | | | **0.01325766** | 1.0 | | | | |
| Mean | 0.0 | 0.2 | | | 0.2 | - | 0.2 | 0.1 |
| SD | 0.0 | 0.2 | | | 0.3 | - | 0.1 | 0.2 |
| Range | 0.000-0.000 | 0.000-0.893 | | | 0.000-0.893 | - | 0.000-0.382 | 0.000-0.893 |

**Supplementary Table 6**. The mean, standard deviation, and range of each of the top 10 tumor features stratified by DNA mismatch repair (MMR) proficient (pMMR) and MMR-deficient (dMMR) status including by the three dMMR subtypes (dMMR-Lynch, dMMR-MLH1me and dMMR-DS) from panel sequenced A) colorectal cancers (CRCs), B) endometrial cancers (ECs) and C) sebaceous skin tumors (SSTs). Statistically significant p-values are highlighted in **bold**. dMMR-Lynch, Lynch syndrome; dMMR-MLH1me, *MLH1* methylated; dMMR-DS, double somatic MMR mutation; SD, standard deviation; INDEL, insertions and deletions; TMS, tumor mutational signature; SBS, single base substitutions; ID, small insertions, and deletions; TMB, tumor mutation burden (mutations / mega base); * heteroscedastic, two-tailed t-test.
